# Radiomics Analysis of Clinical Myocardial Perfusion Stress SPECT Images to Identify Coronary Artery Calcification

**DOI:** 10.1101/2021.01.29.21250803

**Authors:** Saeed Ashrafinia, Pejman Dalaie, Mohammad Salehi Sadaghiani, Thomas H. Schindler, Martin G. Pomper, Arman Rahmim

## Abstract

**Purpose:** Myocardial perfusion stress SPECT (MPSS) is an established diagnostic test for patients suspected with coronary artery disease (CAD). Meanwhile, coronary artery calcification (CAC) scoring obtained from diagnostic CT is a highly specific test, offering incremental diagnostic information in identifying patients with significant CAD yet normal MPSS scans. However, after decades of wide utilization of MPSS, CAC is not commonly reimbursed (e.g. by the CMS), nor widely deployed in community settings. We aimed to perform radiomics analysis of normal MPSS scans to investigate the potential to predict the CAC score.

**Methods:** We collected data from 428 patients with normal (non-ischemic) MPSS (^99m^Tc-Sestamibi; consensus reading). A nuclear medicine physician verified iteratively reconstructed images (attenuation-corrected) to be free from fixed perfusion defects and artifactual attenuation. 3D images were automatically segmented into 4 regions of interest (ROIs), including myocardium and 3 vascular segments (LAD-LCX-RCA). We used our software package, standardized environment for radiomics analysis (SERA), to extract 487 radiomic features in compliance with the image biomarker standardization initiative (IBSI). Isotropic cubic voxels were discretized using fixed bin-number discretization (8 schemes). We first performed blind-to-outcome feature selection focusing on a priori usefulness, dynamic range, and redundancy of features. Subsequently, we performed univariate and multivariate machine learning analyses to predict CAC scores from i) selected radiomic features, ii) 10 clinical features, iii) combined radiomics + clinical features. Univariate analysis invoked Spearman correlation with Benjamini-Hotchberg false-discovery correction. The multivariate analysis incorporated stepwise linear regression, where we randomly selected a 15% test set and divided the other 85% of data into 70% training and 30% validation sets. Training started from a constant (intercept) model, iteratively adding/removing features (stepwise regression), invoking Akaike information criterion (AIC) to discourage overfitting. Validation was run similarly, except that the training output model was used as the initial model. We randomized training/validation sets 20 times, selecting the best model using log-likelihood for evaluation in the test set. Assessment in the test set was performed thoroughly by running the entire operation 50 times, subsequently employing Fisher’s method to verify the significance of independent tests.

**Results:** Unsupervised feature selection significantly reduced 8×487 features to 56. In univariate analysis, no feature survived FDR to directly correlate with CAC scores. Applying Fisher’s method to the multivariate regression results demonstrated combining radiomics with the clinical features to enhance the significance of the prediction model across all cardiac segments. The median absolute Pearson’s coefficient values / p-values for the three feature-pools (radiomics, clinical, combined) were: (0.15, 0.38, 0.41)/(0.1, 0.001, 0.0006) for myocardium, (0.24, 0.35, 0.41)/(0.05, 0.004, 0.0007) for LAD, (0.07, 0.24, 0.28)/(0.4, 0.06, 0.02), for LCX, and (0.06, 0.16, 0.24)/(0.4, 0.2, 0.05) for RCA, demonstrating consistently enhanced correlation and significance for combined radiomics and clinical features across all cardiac segments.

**Conclusions:** Our standardized and statistically robust multivariate analysis demonstrated significant prediction of the CAC score for all cardiac segments when combining MPSS radiomic features with clinical features, suggesting radiomics analysis can add diagnostic or prognostic value to standard MPSS for wide clinical usage.

## 1 Introduction

Myocardial perfusion (MP) SPECT is established for non-invasive evaluations of patients suspected with coronary artery disease (CAD) [1, 2]. It is the most widely-used technique of nuclear cardiology, and its purpose is to assess the adequacy of blood flow to the myocardium [3]. MP imaging can be performed with either planar or tomographic techniques [3, 4], while nowadays, tomographic MP imaging through SPECT scanners has become increasingly more accessible, affordable, and prevalent.

MP stress SPECT (MPSS) has an established pathophysiologic basis with radiotracers capturing blood flow. If a patient with CAD is at rest, typically, blood flow through a diseased coronary artery (e.g. narrowed through plaque build-up) is not decreased until coronary stenosis exceeds 90% of the artery. On the other hand, coronary reserve, i.e., the ability to increase coronary blood flow in case of increased metabolic demand, is reduced if coronary stenosis exceeds 50% [5, 6]. As a result, patients who suffer from CAD may depict homogenous uptake of myocardial blood flow even in the presence of a severely narrowed coronary artery. But the same degree of narrowing can result in reduced flow reserve when the heart is stressed under exercise, resulting in inhomogeneity of regional MP. Such inhomogeneity can be captured using radiotracers that are distributed in the body in proportion to myocardial blood flow [3].

Large prospective studies have shown that coronary artery calcification (CAC) scoring, performed based on diagnostic CT images, is associated with the risk of future cardiovascular events [7-10]. Studies have shown that noninvasive tests for CAD including electrocardiogram (ECG), ultrasound imaging, and even MP SPECT scan, which are used frequently in assessment and diagnosis of cardiac patients, have been of limited value to detect this calcification due to their low sensitivity [11]. At least 25% of patients that experience non-fatal acute myocardial infarction or sudden death have not presented symptoms [12], and it is necessary to identify asymptomatic individuals at greater risk of future cardiovascular events for preventive measures.

The main CAC scoring protocol involves the Agatston method [13] which is often used in clinical practice. The score is calculated for each of the main arteries of the heart, namely the left anterior descending (LAD), the right coronary artery (RCA), and the left circumference (LCX), as well as the left main (LM). This calculation, despite being relatively straight-forward, requires special software, and the cost associated with its licensing requirements might be another hurdle to its widespread application in small radiology centers.

CAC is a highly specific marker of coronary atherosclerosis, and higher CAC scores are associated with increased plaque burden and increased cardiovascular risk [14, 15]. Previous studies demonstrated that a considerable number of stenoses do not result in abnormal perfusion on MP imaging [16, 17], which is why in our work we set the inclusion criteria of ‘non-ischemic normal’ MP stress scans. Furthermore, the CAC score is shown to offer incremental diagnostic information over MP SPECT for identifying patients with significant CAD and *negative* MP imaging results [18]. Unlike MPS, the CAC test is *not* commonly reimbursed, e.g. by the Center for Medicare & Medicaid Services (CMS), while it is known to improve risk stratification in asymptomatic individuals [14, 16], and is not commonly deployed especially in community settings. Moreover, CAC calculation requires sophisticated software and trained radiologists. It is included in the diagnosis package of CAD patients in large institutions but is not readily available in community settings. It is worth mentioning that standalone SPECT cameras (no CT) comprise ∼80% of SPECT market share, further motivating our effort to enable added value to SPECT-only images [19].

In the present work, we aim to assess the added clinical utility of routine clinical myocardial perfusion (MP) SPECT imaging through advanced radiomics analysis. We hypothesize that identification of mild heterogeneities via radiomic analysis may enable identification of subclinical CAD that would carry important diagnostic and prognostic information. We aim to evaluate our hypothesis that MP SPECT radiomic features extracted from clinically normal (non-ischemic) MP SPECT scans correlate with coronary artery calcification (CAC) as extracted from CT imaging.

The field of radiomics transforms digitally encrypted medical images that contain information regarding tissue pathophysiology into mineable high-dimensional data [20, 21]. It hypothesizes that different phenotypic characteristics such as intra- and inter-tissue uptake heterogeneity can be quantified as features (‘radiomic features’) through advanced image processing and computer vision techniques [20]. The information is harnessed through image processing and quantitative image analyses [22], and can be leveraged via clinical decision support systems to improve decision making and personalized medicine [23]. In this study, we developed a pipeline to evaluate various classes of standardized radiomic features of clinical MP SPECT images. Radiomics is a relatively young discipline and has experienced relatively fast growth, yet it remains to be translated to routine clinical practice. This may be due to the *low reproducibility* of most current studies [24]. Radiomics has a complex workflow involving many steps that often suffers from incomplete reporting of methodologic information. Consequently, few radiomics studies available in the current literature can be readily reproduced from start to end [24]. Another major issue is the relatively small number of images in radiomics research datasets that may induce overfitting and high false-positive rates. This further worsens with the tendency to report overly-optimistic results [24].

Guidelines and protocols are available for quality control measures in nuclear medicine imaging to standardize patient preparation, dose production and administration, image acquisition, image reconstruction, intensity normalization, etc., such that the absolute intensity values are interchangeable in multicenter studies [25]. Nonetheless, the methodology to prepare the image and calculate radiomic features is also subject to variability, showing a crucial need for standardization [26-28]. Several studies have shown the importance of robust and standardized protocols to enable reliable quantification of heterogeneity with textural features. They demonstrated an important need to standardize the computation methods due to the complexity of the radiomics workflow [24-26, 29]. An effort initiated in 2016, now comprising 50+ researchers from 25+ universities and cancer centers, including our group, has formed the image biomarker standardization initiative (IBSI) [30, 31]. IBSI has aimed to standardize feature computation and image preprocessing phases of the radiomics workflow to ensure its reproducibility.

Radiomics has witnessed significant activity, especially in oncologic MRI, CT, and PET [26, 32-35]. Yet it has *not* been thoroughly assessed in SPECT and/or cardiac imaging, partially due to the lower spatial resolution that may appear less likely to provide valuable texture and heterogeneity information. However, our group has successfully demonstrated the potential use of radiomics in brain SPECT [36, 37]. At the same time, cardiac SPECT radiomics remains to be thoroughly explored. Moreover, the prevalence of these scans is significantly higher compared to PET exams, enabling collections of a higher volume of data for such data-oriented MP SPECT research [38].

In the present work, we aim to investigate the relationship between MP SPECT radiomic features and the CAC scores, utilizing a large dataset, and performing standardized radiomics analysis. In doing so, we hope to consider a new possibility, namely to use clinical MPSS scans for additional assessment of CAC (Figure 1). This has important implications, given that, as mentioned above, CAC assessment is not commonly performed nor reimbursed in a wide community setting, and as such, our proposed framework holds promise for new added usage and value for routine MPSS imaging. In what follows, we describe our methods, following by results, discussion, and conclusion.

**Figure 1.**
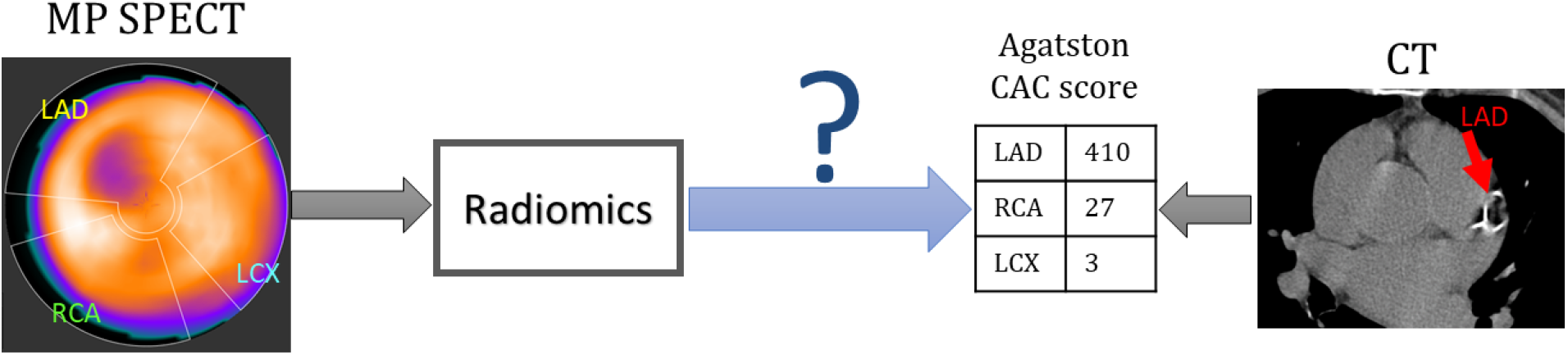
Diagram of the problem addressed in this paper: using radiomics of stress MPSS to predict CAC scores from CT.

## 2 Methods

Our efforts consist of 4 primary steps, elaborated in the next subsections: (i) We start by identification of patients with *normal* MPSS tests and CAC CT, following by (ii) image segmentation. Next, (iii) we extract radiomic features, followed by the elimination of non-reproducible and redundant features (feature selection). Finally, (iv) we use machine learning techniques, specifically multivariate regression, to derive and validate measures for correlation against CAC information from MPSS image radiomics.

### 2.1 Patient Collection

After obtaining approval from the institutional review board (IRB) at Johns Hopkins University, we searched for patients with stress myocardial perfusion SPECT scans from 2011 until 2015. We investigated around 1,800 reports of patients undergone MPSS, out of which 428 cases were selected. All patients had a CT scan for CAC scoring at the same time as their MP SPECT scan in the PACS database. A nuclear medicine physician investigated the MPSS images to be free from i) image artifacts, ii) overcorrection, and iii) spillover from nearby liver or stomach, deriving detailed CAC score for each of the 4 arteries of the heart using a clinical software.

The dataset consists of images collected from various Siemens®, GE®, and Phillips® scanners, at the Johns Hopkins Hospital throughout the above-mentioned years, while all were reconstructed with an attenuation-corrected iterative reconstruction (AC-IR) algorithm and with a consistent voxel size of 4.8 mm. According to quality factors of radiomics research, it is an important characteristic of a study to have imaging acquisition protocols that are “well described and ideally similar across patients”, and involve “methodologic steps taken to incorporate only images of sufficient quality” [24].

We recorded various parameters for each patient, including basic information (age, sex, race, height, and weight at scan time), clinical history (smoking, diabetes, hypertension, hyperlipidemia, and family history of cardiac disease), and scan info (voxel size, slice thickness).

### 2.2 Image segmentation

The study involves three different layers of segmentation as applied to MPSS images: i) total myocardium, ii) 3 vascular segments, and iii) 17 polar segments (feature evaluation and statistical analysis were performed over these segmentations). The segmentation methods are presented in Figure 2. We selected two different methods for vascular segmentation as both are widely used in the clinic. Unlike the 17 polar segments that span the entire heart, the 3 vascular segments method comprise of three more stringent segments with gaps between them as can be observed from Figure 2.

**Figure 2.**
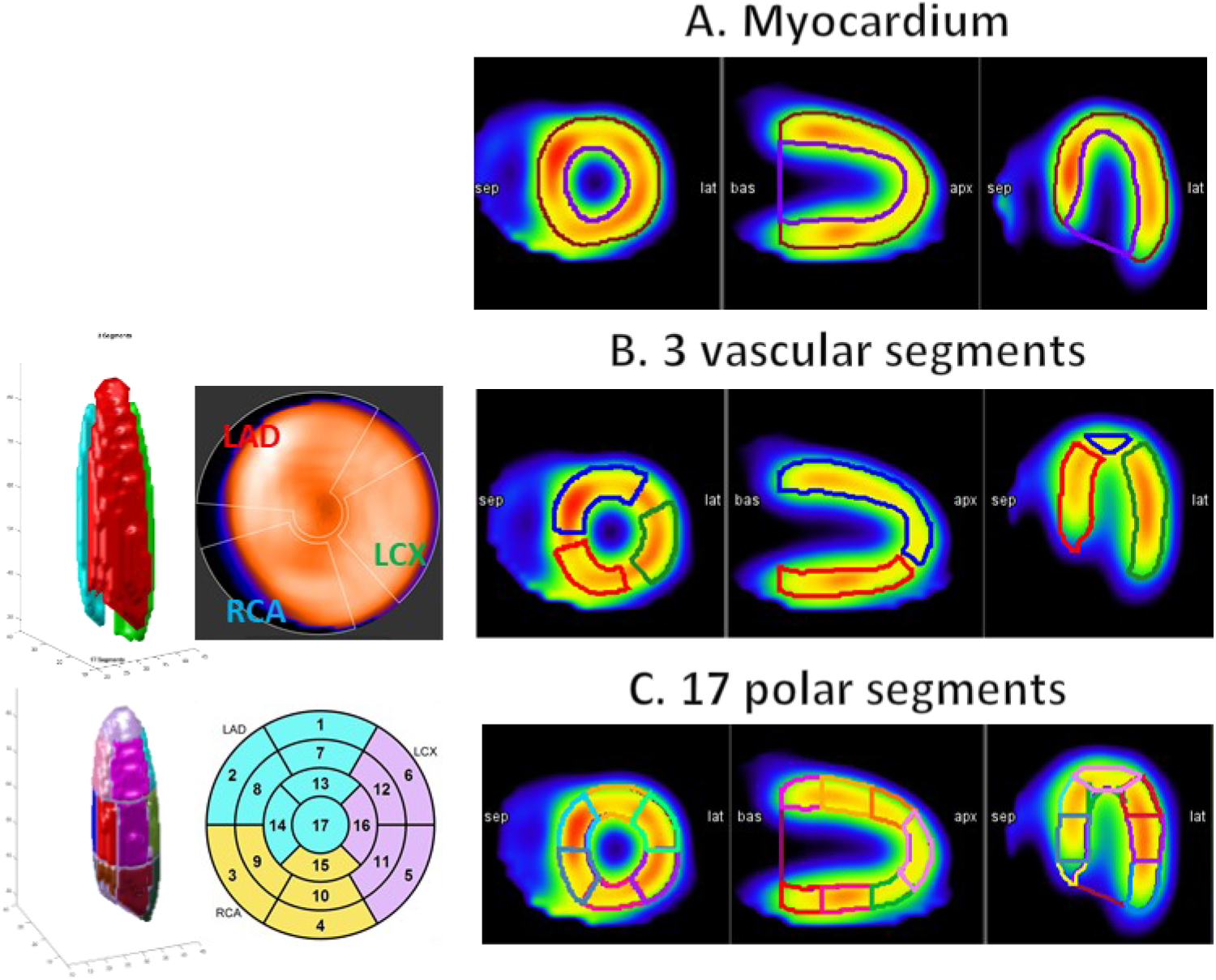
Three methods of segmentation used in our study. A) the myocardium segment. B) 3 vascular segments of the heart (LAD, RCA, and LCX), and C) subsets of 17 polar segments of the heart grouped into LAD, LCX, and RCA.

We used MIM software^®^ and developed a workflow that automatically draws 3D contours over 21 regions of the heart, namely: endocardium, epicardium, 3 vascular segments (Figure 2B), and 17 polar segments (Figure 2C). The workflow generated the myocardium segment using epi- and endocardium segments. The automated segmentation was supervised by a radiologist to assure the correct placement of the contours over the corresponding segments. Our workflow then exported the 3D SPECT image and its 22 contours as 3D MATLAB^®^ matrices, which were subsequently used for analysis.

### 2.3 Radiomics Framework

We used our developed standardized environment for radiomics analysis (SERA) software package, which we have made publicly available (see ‘code availability’), deriving radiomic features that are standardized and reproducible, consistent with guidelines by the Imaging Biomarker Standardization Initiative (IBSI) [30, 31]. IBSI is a global initiative consisting of >25 universities and cancer centers, in which our group is an active participant. SERA calculates up to 487 standardized radiomic features aiming to standardize the preprocessing and feature evaluation phases, and to meet standards by the IBSI, in order to contribute to reproducible research [39]. Some of the radiomic features by SERA have also been validated in another standardization study [40]. Section S.2 in Supplementary Materials includes a brief introduction to SERA and its configuration, in addition to a spreadsheet of computed features from IBSI benchmark phantoms.

Images produced for MP SPECT scans have an arbitrary unit – counts – that do not quantitatively relate to biological phenomena. Therefore, we ought to use fixed bin number discretization of the counts. We considered and investigated 8 different grey-levels (GL) discretization, specifically using 4, 8, 16, 32, 64, 128, 256, and 512 bins. All images in our dataset were reconstructed into 3D images with identical isotropic voxel sizes of 4.8×4.8×4.8 mm^3^; thus, no resampling and interpolation were needed. We did not perform any GL rounding or re-segmentation. The framework was then ready to calculate 487 features for 8 GLs over 7 different segmentations of the heart.

### 2.4 Statistical Analysis

We used multiple analyses to eliminate non-useful features, including features that are identical, non-robust, and redundant. We performed a multistep feature selection to significantly reduce the size of our feature-space of 487×8 features. This process was performed completely independent of outcome (e.g. CAC score, etc.). The selected feature set was subsequently passed on to univariate and multivariate analysis schemes to correlate with CAC measures. We also accounted for false-discovery by employing false-discovery rate (FDR) correction, specifically using the Benjamini-Hochberg method [41].

#### 2.4.1 Feature Selection

SERA calculates all 487 features as defined in the IBSI documentation that are from 11 main feature categories, namely statistical, morphological, local intensity, histogram, intensity histogram, GL cooccurrence, GL run length, GL size-zone, GL distance-zone, neighboring grey tone difference, neighborhood grey tone difference, and neighboring grey level dependence. All these features were initially considered and were calculated for 8 different GLs. In this section, we aim to systematically narrow down this large feature set and arrive at a smaller set of meaningful, robust, non-redundant, and reproducible features for further investigation of their predictive value, discouraging overfitting. Our feature selection phase can be generally divided into i) pre- and ii) post-feature-computation steps, as explained next. Following feature selection, we discuss how to narrow down to an optimum discretization level.

##### Pre-computation feature selection

In the first step, we eliminate irrelevant feature families based on the nature of our dataset and our *a priori* knowledge about what each feature captures and its usefulness.

Removing 2D and 2.5D feature families: Our dataset originally consists of images with isotropic voxels. As such, there is no additional information provided to us from 2D or 2.5D feature families. These feature families are distinct only when slice thickness (i.e. voxel size in z dimension) is different from the voxel size in x and y dimensions. In that case, resizing and interpolating the images to isotropic voxel sizes may have resulted in modification of the original voxel distribution, causing possible loss or modification of data. In any case, the following feature families were eliminated: 2D and 2.5D GLCM (25 features) and GLRLM (16 features) (both merged and averaged), 2D and 2.5D GLSZM (16 features), GLDZM (16 features), NGTDM (5 features), and (17 features). This removed 272 features, narrowing down our feature space to 215.

Removing useless feature families: MP SPECT images have voxels with arbitrary units (they are typically not quantitative, unlike PET or some SPECT imaging applications). Therefore, any feature that conveys information regarding the exact intensity values of the original ROI is not considered meaningful. As such, intensity-based features (18 features) and local intensity features (2 features) were excluded. Furthermore, the 7 segments were created by an automatic segmentation procedure that generates ROIs with similar shapes (all registered to the same reference space). As such, the shape of the segments does not carry any differentiating information, and we are interested mainly in the heterogeneity caused by voxel intensity variations which carry information about blood flow in different heart segments. As such, morphological features (29 features) were excluded. At the end of this step, we were left with 166 features out of 487, eliminating the majority.

##### Post-computation feature selection

Following feature computation, we run the following multistep post-calculation feature selection, to eliminate redundant information. This includes the removal of (i) feature with identical values, (ii) features in ‘families’ with more than one variety (such as GLCM 3D averaged vs. GLCM 3D combined) using the Spearman rank correlation between each feature and all other features, (iii) features with a low dynamic range (percent variance of the features (variance/mean)) less than 10^−5^, and (iv) highly-correlated features having a Spearman correlation coefficient |*ρ*| ≥ 0.95 as suggested in the literature [42].

##### Selecting the best GL post-computation

We performed the feature computation for 8 different GLs intending to select the most robust GL for our analysis and to eliminate running analyses on all 8 of them. Hence, we followed a blind-to-outcome procedure using the dynamic range of calculated features across all 8 GLs to narrow down our GLs and selected one that demonstrates wider variation and less redundant values across all patients with more potential information compared to other GLs.

#### 2.4.2 Outcome Prediction

Following the above-mentioned feature selection steps, we performed outcome prediction specifically to predict the CAC score of each region of the heart from radiomic features extracted from the same region of the MPSS image. We performed two analyses: univariate and multivariate.

##### Univariate analysis

We started by investigating whether our selected radiomic features (previous section) directly correlate with the CAC score. We adopted two approaches. In the first approach, the CAC score with a continuous scale was utilized. In the second approach, we discretized CAC scores of each region of the heart based on a popular 5-scale clinical stratification criteria (shown in Figure 4) [16]. Spearman rank correlations between features of every segment with the CAC score of the same segment were calculated for both CAC approaches (continuous and discrete). We employed the Benjamini-Hochberg false-discovery rate (FDR) correction with q = 0.05 to discourage false discovery.

**Figure 4.**
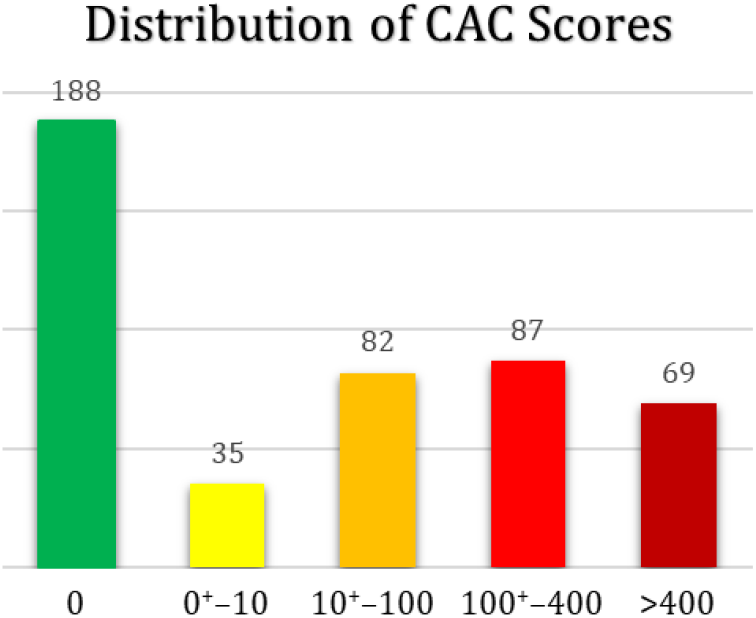
Distribution of patient CAC scores based on widely-used stratification criteria [16].

##### Multivariate analysis

We additionally pursued a multivariate approach to predicting CAC scores. In this subsection, first, we introduce stepwise linear regression, then we describe how we handle feature selection. We then explain how our proposed algorithm analyzes data for robust and reliable analysis, and finally, we run the analysis for three different configurations: i) radiomics features-only, ii) clinical features-only, and iii) radiomic + clinical features, and present the results.

##### Stepwise linear regression

Stepwise regression was used as a systematic method for adding and removing terms from a linear or generalized linear model based on statistical significance in explaining the response variable. The method began with a constant (intersect) model, and then compared the explanatory power of incrementally larger and smaller models, performed by adding or removing terms by stepwise regression and returning the linear model at the end. At every iteration, the function examined a set of available features and added the best ones to the model if an F-test for adding the term results in a *p*-value of *P*_*enter*_, or less. If no terms could be added, it examined the terms currently in the model and removed the worst one if an F-test for removing it had a *p*-value of *P*_*remove*_, or greater. This process was repeated until no terms could be added or removed. The constant term (intercept) was never removed from the model.

##### Feature handling

At each step, the method searched for terms to add to or remove from the model based on the Akaike information criterion (AIC). As an estimate of the relative quality of statistical models for a given dataset, AIC was used to reduce the chance of overfitting as well as underfitting by providing a balance between the goodness of fit and having too many parameters [43].

We can specify the order at which the algorithm starts to add features and later removes them. Instead of an unstructured approach of starting from the arbitrary first feature in the list, we developed a feature selection method to enter those with higher Spearman correlation to the model first. For this purpose, the univariate Spearman rank-correlation between each feature in the *training* set and the outcome (CAC score of the same segment) was calculated. The Spearman correlation coefficients and their corresponding *p*-values were recorded. Then, features with *p*-values smaller than 0.3 were selected and sorted into descending order, based on the value of their Spearman correlation. The input dataset was then rearranged based on this subset of Spearman correlation-sorted features to enter features with the highest correlation to the stepwise algorithm first.

##### Training/cross-validation/testing setup

A flowchart of the workflow is depicted in Figure 3. The following procedures were performed for each of the cardiac segments separately. First, randomly-selected 15% of the data was set aside as the “independent test set”. This set was not used until the very end as an independent assessment. Then, the following procedure was performed 20 times: the remaining 85% of the data (‘training + dev set) was randomly divided into training and cross-validation sets with 75%/25% ratios.

**Figure 3.**
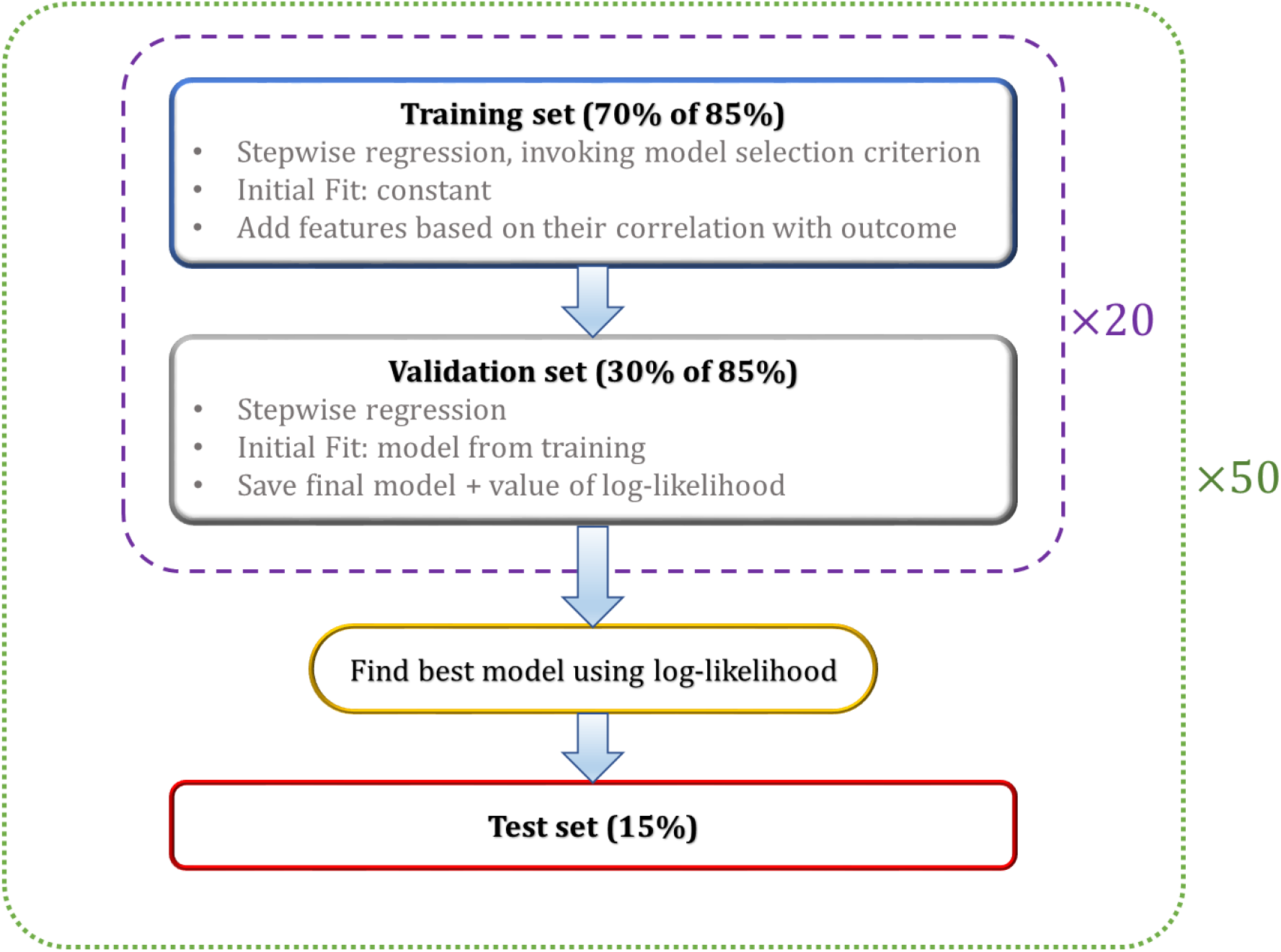
A simplistic flowchart of the algorithm.

We subsequently perform stepwise linear regression first on the training set. We set *P*_*enter*_ as 0.05 and *P*_*remove*_ as 0.2. Following training, we performed cross-validation using the dev set. Cross-validation aims to reduce overfitting to the training set. The cross-validation algorithm is configured the same as training, except for the training algorithm the initial fit was a constant (intercept), whereas for cross-validation the initial fit is the output fit from the training dataset. During the above steps, we recorded the model, including the set of features remaining in it, the value of the log-likelihood, *p*-value, and the final AIC.

The model fit is typically comprised of several features that survive the stepwise algorithm, and it might be possible that only the intercept term survives. If by coincidence the best model consists of only the intercept term, we skip that and choose the best fit with more than one term.

Following the above procedure, we select the model with the highest AIC in the 20 runs as the winning model to evaluate and run on the independent test set—blind to the entire operation. To assess prediction performance, Pearson correlation was used to assess the relationship between the two distributions (prediction vs. actual), and subsequently recorded the correlation coefficients (*ρ*) and their corresponding *p*-values. The above operation was performed for each of the segmented lesions of the heart separately.

Although the test set was not utilized during training/validation, to assure independent and blind-to-training assessment, the results might still be biased to a specific randomly selected test set chosen. To further mitigate such a bias, we took an extra step and performed the above-mentioned operation 50 times. That is, randomly shuffling and dividing the dataset into “training + dev” and “test” sets 50 times, then running the stepwise algorithm 20 times over the “training + dev” set. We subsequently performed 50 predictions on 50 independent test sets, providing us 50 best regression fits and their *p*-values, which we subsequently used to derive our conclusion.

Three configurations of input parameters: We performed the above analysis three times: A) with radiomic features only (imaging), B) with clinical features (non-imaging), and C) with both radiomics and clinical features. The 10 clinical features employed were: i) sex, ii) race, iii) age, iv) smoking, v) diabetes, vi) hypertension, vii) hyperlipidemia, viii) family history of cardiac disease, ix) body-mass index (BMI), and x) left ventricle ejection fraction (LVEF). We also assured that a certain subset of clinical features such as sex, race, diabetes, etc. was treated as “categorical” variables, as opposed to continuous, by the algorithm.

##### Assess the significance of results

We used Fisher’s method for combining the p-values, and the chi-squared distribution test to determine significance after running the 50 random trials. Under the null hypothesis, Fisher’s measure computed as

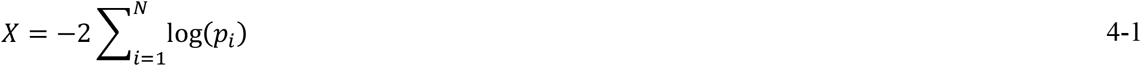

follows a chi-squared distribution with a degree of freedom of 2 × *N*, where *N* is the total number of runs [44] (in our case, *N* = 50). Comparing the value of *X* to the appropriate chi-squared distribution can determine whether the sample is significant. Assuming a significant level of 0.01, the value of the chi-squared distribution for degree-of-freedom of 2×50=100 is 135.81. Section S.3 in Supplementary Materials includes the Matlab code used to run the multivariate analysis workflow comprising the steps detailed in this section to help other researchers reproduce this study, which includes stepwise linear regression adopting AIC followed by Fisher’s method to determine significance.

## 3 Results

### 3.1 Analysis of dataset statistics

The dataset was comprised of 229 female (49.7%) and 232 male (50.3%) subjects. Distributions of patient age, height, weight, and body mass index based on sex is depicted in **Error! Reference source not found**.. We observe a relatively close distribution of age between males and females. Our dataset consisted of 428 normal scans. In our dataset, stress was induced on 274 patients by injection of a vasodilator (264 Adenosine, 7 Dipyridamole, 2 Regadenoson), and on 186 patients via treadmill exercise based on the Bruce protocol [45]. **Error! Reference source not found**. shows the distribution of LVEF for all patients. The ejection fraction compares the amount of blood in the heart to the amount of blood pumped out. It helps to describe how well the heart is pumping blood to the body. The ejection fraction of a normal heart is between 50% to 70%. A higher LVEF may indicate a heart condition such as hypertrophic cardiomyopathy [46, 47]. Other clinical factors are detailed in Table 1.

**Table 1.** Distribution of patients’ morphology in this study

| Factor | Attribute | No. of Cases | % of Total |
| --- | --- | --- | --- |
| **Smoking** |  |  |  |
|  | Non Smoker | 220 | 47.7 |
|  | Current Smoker | 137 | 29.7 |
|  | Previous Smoker | 104 | 22.6 |
| **Diabetes** |  |  |  |
|  | No | 313 | 67.9 |
|  | Yes | 148 | 32.1 |
| **Hypertension** |  |  |  |
|  | No | 138 | 29.9 |
|  | Yes | 323 | 70.1 |
| **Hyperlipidemia** |  |  |  |
|  | No | 243 | 52.7 |
|  | Yes | 218 | 47.3 |
| **Family History of CAD** |  |  |  |
|  | No | 283 | 61.4 |
|  | Yes | 178 | 38.6 |

A well-known stratification method presented by Berman, *et al*., divides CAC scores into five categories of 0, 0^+^ to 10, 10^+^ to 100, 100^+^ to 400, and > 400[16]. The distribution of our patients into these five categories is presented in Figure 4.

The plot in Figure 4 shows the distribution for all patients. Patients with normal MP stress tests were having 178, 32, 77, 80, 38, and 23 cases in each of the five bins of Figure 4. This shows 58.4% of the patients with normal MP stress scan had a non-zero CAC score, out of which 33% had a CAC score <u>≥</u>100. Studies have shown that MP ischemia is rare in patients with CAC<100, whereas in <u>patients with CAC≥100, the chance of myocardial ischemia increases progressively</u> [16]. Also, <u>one-third of patients with normal MPSS had a CAC score ≥100</u>. As previously mentioned, CAC scoring is known for its high specificity and very small false positives, suggesting that assessment of atherosclerosis burden by CAC scoring may be useful in finding CAD when MPSS fails to report. It also underlies our motivation to develop and study a radiomics-based scheme to extract CAC information directly from MPSS scans.

### 3.2 Feature Selection Results

The result of the pre-feature calculation was discussed in the Methods section. Below we present the results of our post-feature calculation feature selection steps.

We searched for features with identical values across all patients for further exclusion. In our dataset, 4 features had identical values across all patients: histogram minimum, maximum, and range, and NGLCM dependence count percentage. We now arrive at 162 features.

In the next step, we calculate the Spearman rank correlation between each feature and all other features to explore the relationship of the features with respect to each other and find redundant and highly correlated features. At this step, we had one subtype of every higher-order feature class (i.e. only 3D, after excluding 2D and 2.5D) except for GLCM and GLRLM, each remaining with two 3D subtypes: 3D merged, and 3D averaged. We investigated the correlation between each variety of higher-order 3D calculations, i.e. 3D GLCM averaged vs. merged, and 3D GLRLM averaged vs. merged. Figure 5 shows a heatmap of their correlation. In the diagonal of both heatmaps in Figure 5, we observe very high Spearman correlation values (between 0.98 to 1) between all the same features within the two feature families, i.e. GLCM-averaged entropy vs. 3D GLCM-merged entropy, etc., indicating the redundancy of features calculated in two varieties (merged vs. averaged). Let us *S*_{*A*}|{*B*}_ as the Spearman rank correlation between feature families {*A*} and {*B*}. We subtracted *S*_{3D GLCM-averaged}|{All feature families except 3D GLCM-merged}_ from *S*_{3D GLCM-merged}|{All feature families except 3D GLCM-averaged}_, and did the same for GLRLM, and observed it yields values very close to zero, which further indicates that using one variety vs. the other does not add additional information to our analysis, suggesting the exclusion of one variety from both GLCM and GLRLM. Subsequently, to decide which of the two varieties to exclude, we calculated the range of features in both varieties and removed the one with a smaller range, which yields to exclusion of the 3D-merged of both categories and keeping 3D GLCM-averaged and 3D GLRLM-averaged. This further reduced the number of features down to 121. This observation is also consistent with findings in [48], where the authors reported merged features with tighter distribution in a smaller range, and subsequently remove, them from the rest of their study.

**Figure 5.**
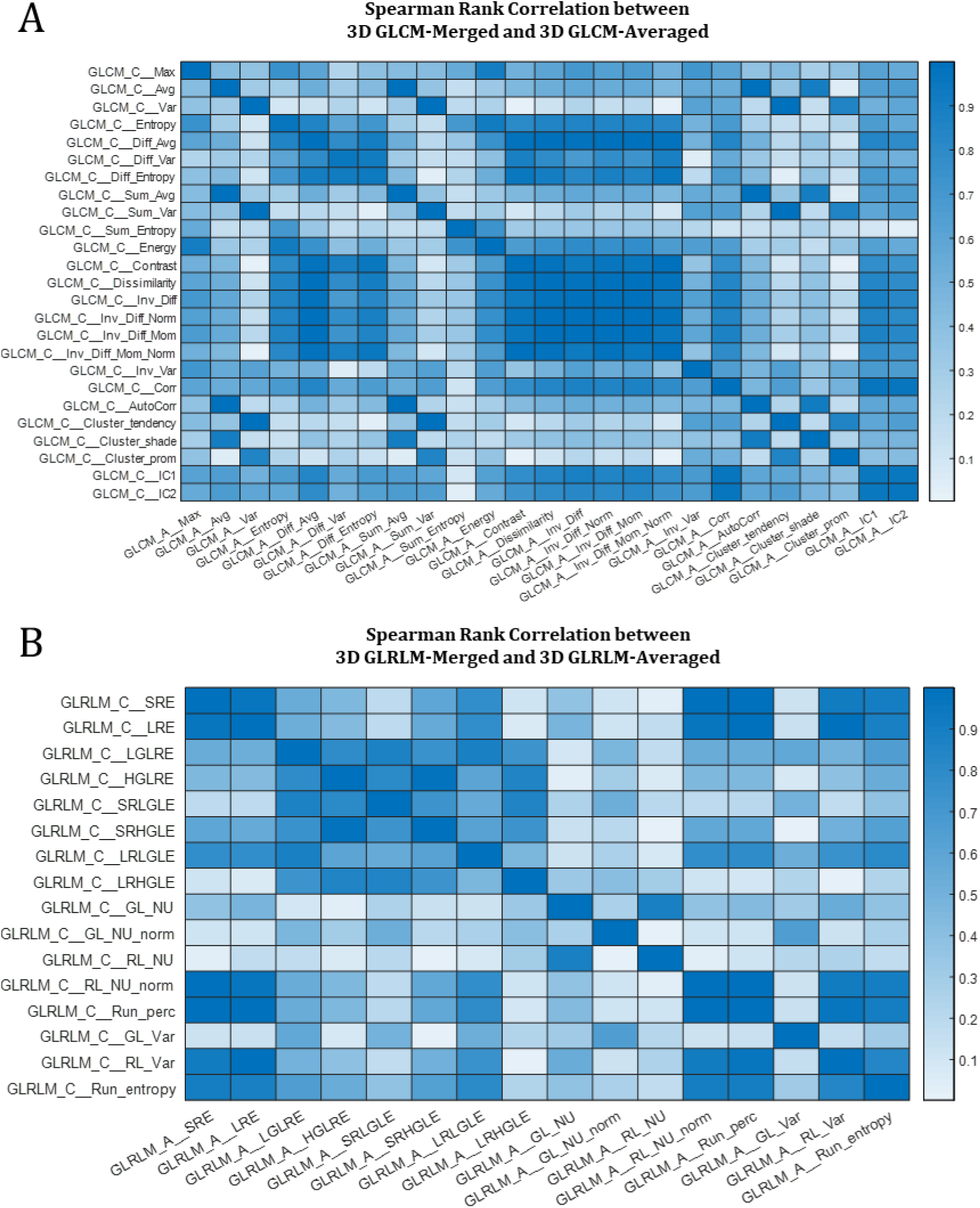
Heatmaps of Spearman rank correlation between A) 3D GLCM-averaged vs. 3D GLCM-merged and B) 3D GLRLM-averaged vs. 3D GLRLM-merged. The diagonal of both plots have values >0.98.

After using Spearman correlation to reduce the feature space at the feature-family level, we move on to investigate the correlation at the feature level. The next set of features to remove is the feature-pair with Spearman rank correlation coefficient of 1, indicating their redundancy. These features included three pairs: i) “3D GLSZM-zone percentage (ZP)” and “3D GLDZM-ZP”, ii) “3D GLSZM-GL nonuniformity (NU) normalized” and “3D GLDZM-GL NU normalized”, and iii) “3D GLSZM-GL NU” and “3D GLDZM-GL NU”, From each pair, we selected the feature with a lower range to exclude that yielded the removing of the GLDZM features from each pair.

Subsequently, we removed features with a very low dynamic range less than 10^−5^, which were five: Histogram-skewness, Histogram-kurtosis, Histogram-min gradient, GLCM-averaged cluster shade, and GLCM-averaged 1^st^ measure of information correlation. Now the dataset has 113 features.

In the last step of this phase, using the Spearman correlation of features with respect to each other calculated earlier, we opt to remove highly correlated features as defined by those having a Spearman correlation coefficient |*ρ*| ≥ 0.95 as suggested in the literature [42]. These feature-pairs are considered to be highly correlated and likely to provide redundant rather than complementary information. We remove these features through the following recursive operation.

We use the heatmap of feature-pair Spearman correlation to find features with |*ρ*| ≥ 0.95. We subsequently record the number of instances a feature fits this criterion. Then, we sort these features based on which feature has more instances of |*ρ*| ≥ 0.95 with others in descending order and call it ℱ^*sorted*^. We then start from the first feature in this set. We denote this first feature by 𝒻^*keep*^, i.e. the feature to keep,and save it to 𝒦 that denotes the set of features we intend to keep. Subsequently, we mark the highly correlated features with 𝒻^*keep*^ and save them to an empty set denoted by ℛ, i.e. for removal. We then loop over each feature inside ℛ and find other highly correlated features with these features and append them to ℛ. Once the procedure is complete, we update ℱ^*sorted*^ by removing 𝒻^*keep*^ and all features inside ℛ. The algorithm then continues recursively with this update ℱ^*sorted*^, letting its first member be 𝒻^*keep*^ and append it to 𝒦, and find features and add them to ℛ for removal. This process continues until ℱ^*sorted*^ becomes empty.

The above algorithm cuts the number of features into half, removing 57 features from 113, yielding 56 features remained that are not highly correlated with each other and are more likely to provide complementary information. Table 2 contains the list of these 56 features.

**Table 2.**
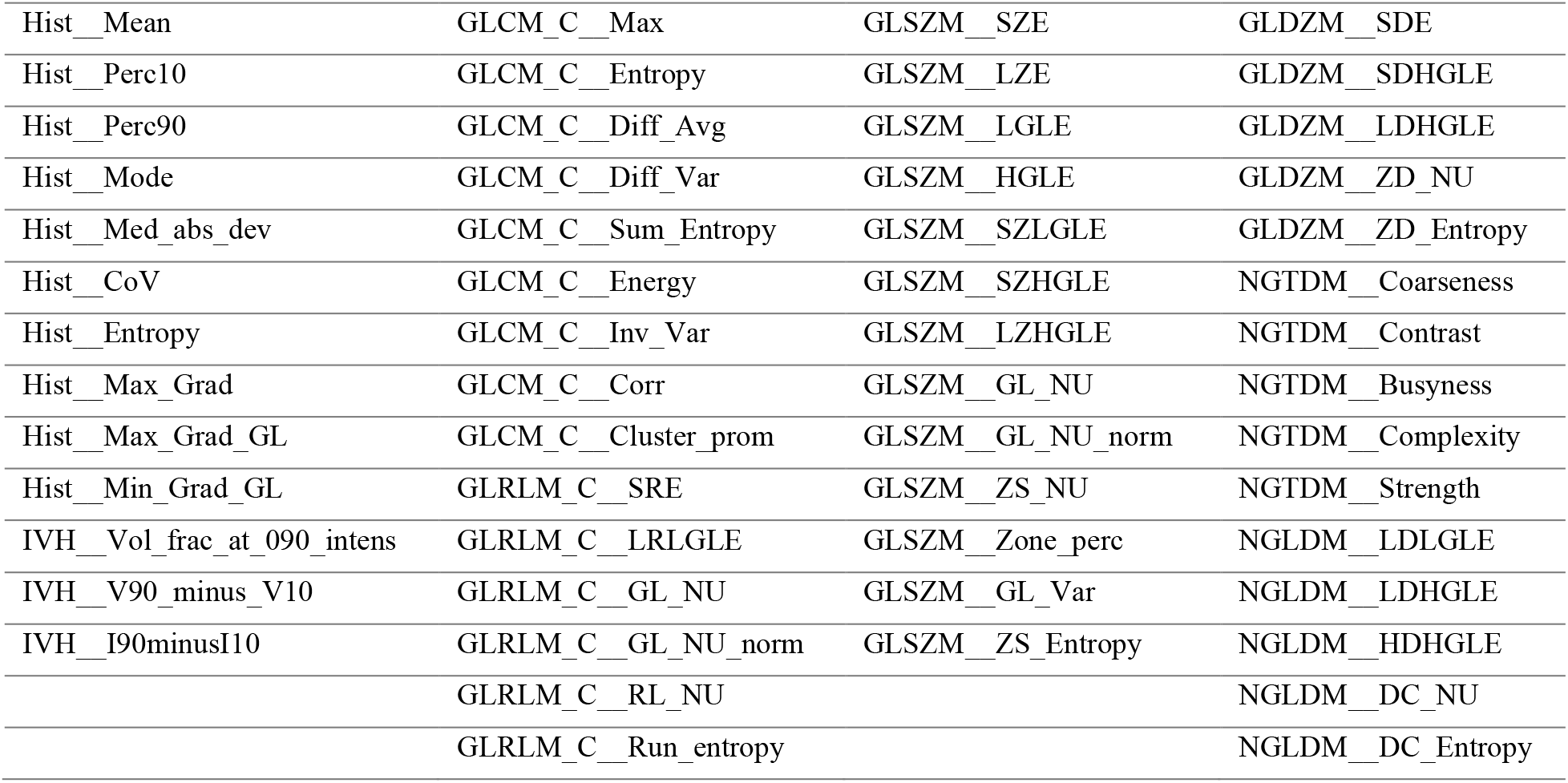
List of 56 robust features from the feature selection phase. This phase was performed blind-to-outcome, thus the set has the potential to be adopted in future MP SPECT studies. Feature acronyms are from the IBSI nomenclature in its guideline [30].

The above procedure reduced the feature set from 487×8 to 56×8 features for 8 GLs. Now we focus on discretization levels to systematically remove non-useful GLs. Firstly, we observe that for the three smallest GLs, the number of identical features is higher than the other five GLs. Furthermore, features with smaller dynamic range increase by 22%, 4%, 29%, and 29% compared to GL=64 or 128. Moreover, the two highest GLs have 11% and 22% more feature-pairs with Spearman correlation ≥ 0.9. Therefore, we can safely remove all GLs except 64 and 128.

The procedure in the previous paragraph could have been performed without the analysis of the range and Spearman rank correlation of features. We can safely remove the first three GLs since the intervals that voxel intensities were discretized into are so large that they do not provide enough opportunity to capture the heterogeneity of a region. On the other hand, the two largest GLs produce so many bins to discretize voxels into that many bins will be empty or just have very few representations in the ROI. For instance, the LAD segment consists of averagely 460 voxels. When it is discretized into 512 GLs, they are actually more bins than voxels, and many bins would be empty or just occur very scarcely. In this case, our higher-order matrices such as GLRLM, GLSZM, GLDZM, etc. in which the number of columns represents different run-lengths, zone sizes, distance zones, etc., would be very long and narrow matrices with very small variability. As a result, these higher GLs should be eliminated, too. Interestingly, this finding is consistent with some previously published studies on radiomics of PET imaging [48, 49].

Finally, out of the remaining two GLs, 64 and 128, we found very similar behavior from both discretization levels in terms of the range of the features and number of feature-pairs with high Spearman correlation. We decided to choose 64 for the rest of this study, because 1) as mentioned GL=128 does not demonstrate different statistical properties, 2) our results in the previous study suggested 64 GLs for the other SPECT study – imaging of renal cell carcinoma with ^99m^Tc, which is the same radiotracer as the one used for MPSS imaging [50], and 3) some previous studies have demonstrated that GL=64 provided higher textural feature reproducibility [51] and robustness [49].

Through the above procedures, we reduced our feature set of 487×8 to 56. One important note is that these features were excluded in a completely unsupervised manner without involvement of clinical outcome (e.g. CAC score, patient survival, etc.). This is an important consideration to make our subsequent efforts statistically sound; further they increase utility of these features for future applications in MP SPECT radiomics.

### 3.3 Outcome prediction results

In this section, we elaborate on our efforts towards outcome prediction using the narrowed down feature set. We also report our negative findings and unsuccessful attempts, as we believe reporting them helps future researchers, and thus, is of scientific value.

#### 3.3.1 Univariate analysis

Figure 6 shows the absolute value of Spearman correlation coefficient values between 56 selected radiomic features and discretized CAC for eight segments, where we can observe the mediocre correlation values. Figure 7 shows their corresponding *p*-values (not FDR corrected in this plot). Following Benjamini-Hochberg FDR correction no feature survives. This emphasizes the difficulty of the task at hand, and that it is necessary to adopt a more sophisticated, multivariate algorithm for regression (for continuous CAC outcome) or classification (for discrete CAC outcome).

**Figure 6.**
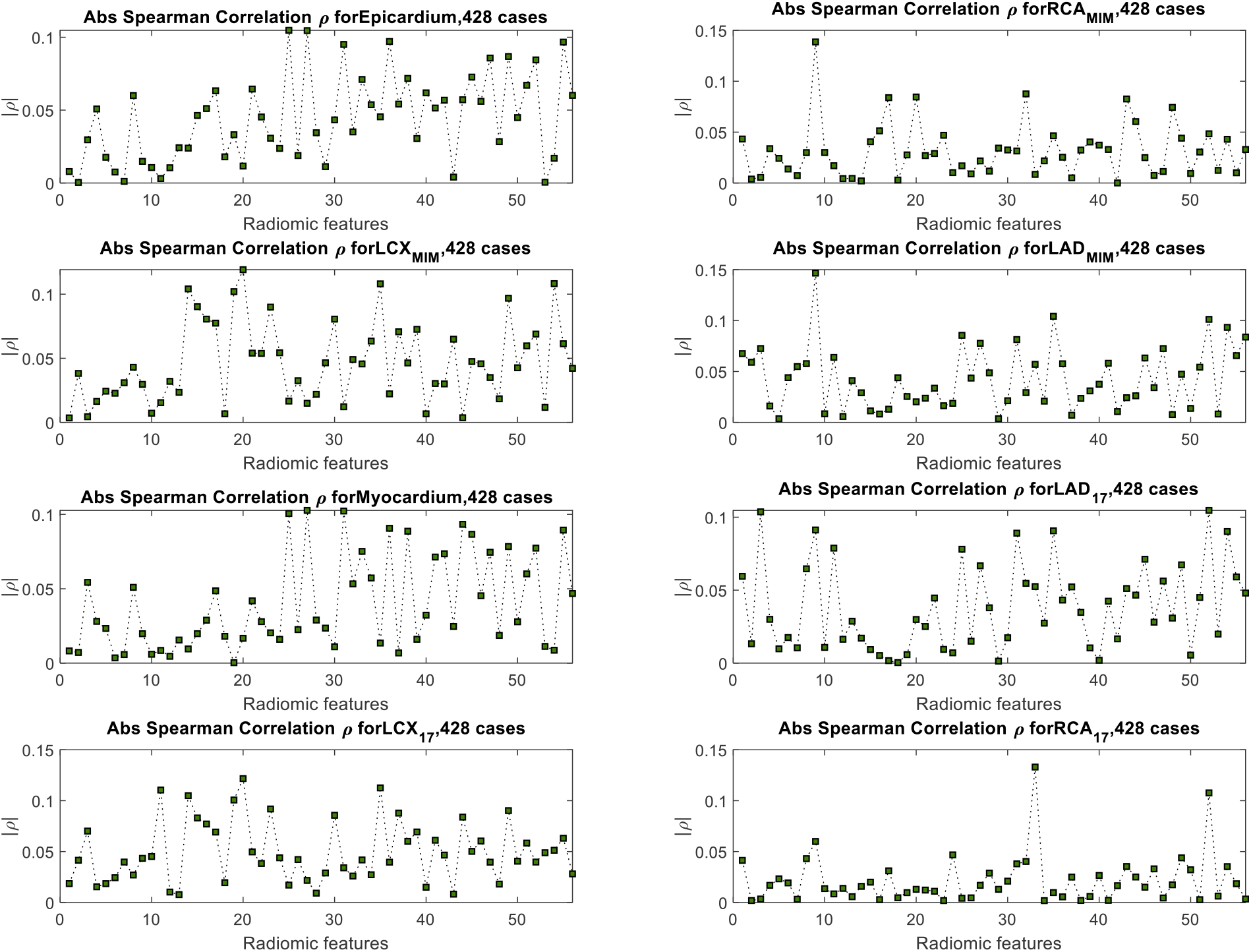
Spearman rank correlation between a selected feature of each segment (56 selected features) and the CAC of that segment. The maximum correlation observed in all plots is 0.15, which is mediocre.

**Figure 7.**
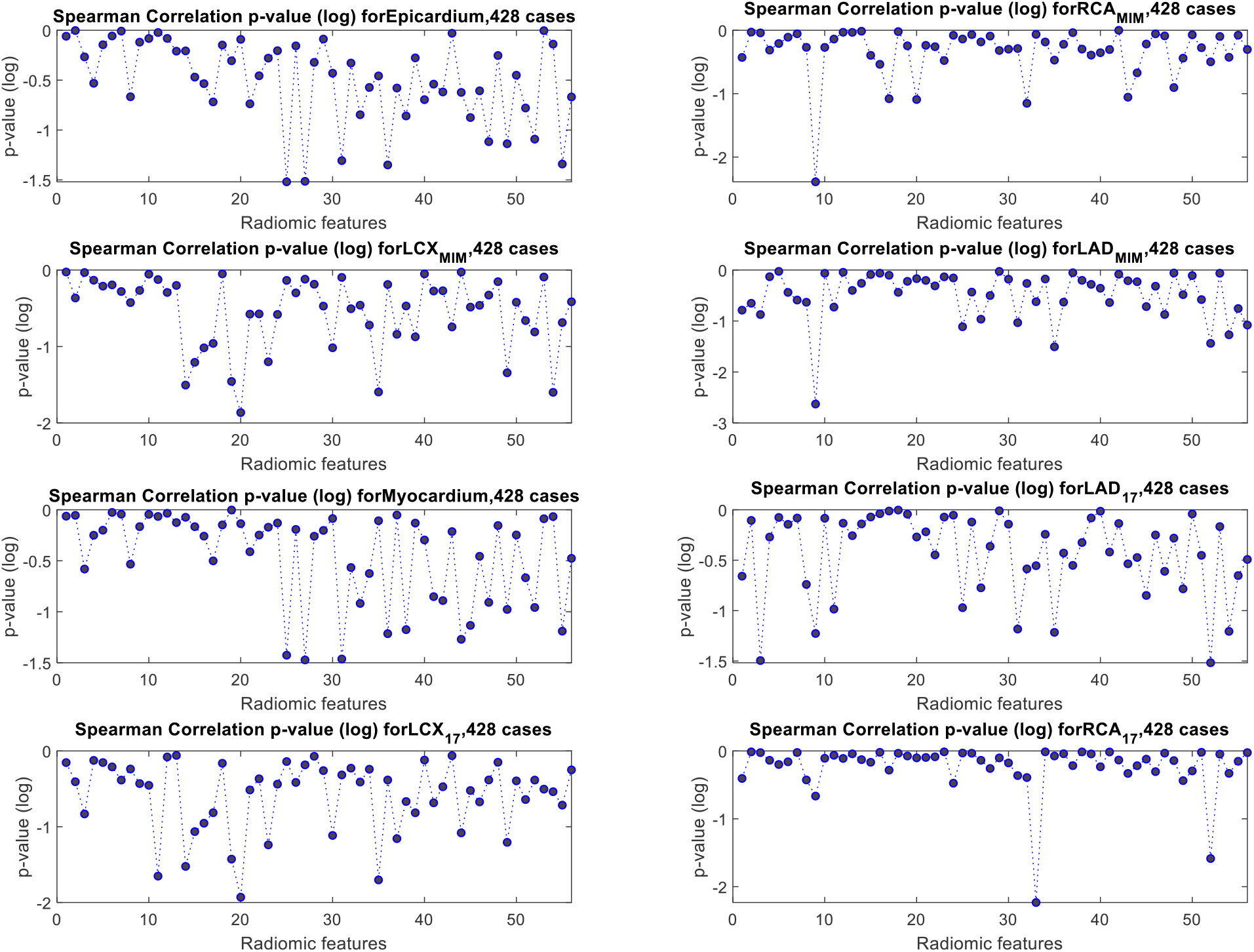
Spearman rank correlation p-value between a selected feature of each segment (56 selected features) and the CAC of that segment.

#### 3.3.2 Multivariate analysis

We observed from Figure 6 that in general, the correlation values between features and CAC scores are relatively low. But despite their low correlation, these selected features had a relatively-higher significance, giving us the hope that a certain multivariate combination of them might actually be predictive and provide significant prediction information. Table 3 shows the result of applying Fisher’s method to the three configurations, where significant results are shown in bold. We observed that radiomic features were unable to yield a significant model for any of the segmentation, and clinical features were able to result in a significant fit for most of the segments. But the combined clinical + radiomic features results in a significant fit across all segments.

**Table 3.** The value of chi-squared distribution for each segment and feature configurations. The value of the chi-squared distribution with a degree-of-freedom of 100 is 135.81, and values above this threshold (shown in bold) are considered significant under the null hypothesis.

|  | RCA<sub>MIM</sub> | LCX<sub>MIM</sub> | LAD<sub>MIM</sub> | Myocardium | LAD<sub>17</sub> | LCX<sub>17</sub> | RCA<sub>17</sub> |
| --- | --- | --- | --- | --- | --- | --- | --- |
| Radiomics | 95.87 | 88.02 | 115.02 | 111.93 | 139.25 | 53.8 | 53.28 |
| Clinical | 84.12 | **153.14** | **253.13** | **294.43** | **253.13** | **153.14** | 84.12 |
| Combined | **174.53** | **194.73** | **348.97** | **341.39** | **326.97** | **189.2** | **141.6** |

Figure 8 shows the distribution of the absolute value of Pearson’s correlation coefficient |*ρ*| for all seven segments. We observe the same pattern across all segments that the combined radiomics + clinical features are more correlated to the CAC scores of that region. Moreover, Figure 9 shows the distribution of *p*-values of the best fit out of the 50 independent runs of the stepwise regression algorithm, each include 20 model fits where the best is selected. This plot shows that adding radiomics to the 10 clinical features will enhance the significance of the regression model and promising a more robust prediction.

**Figure 8.**
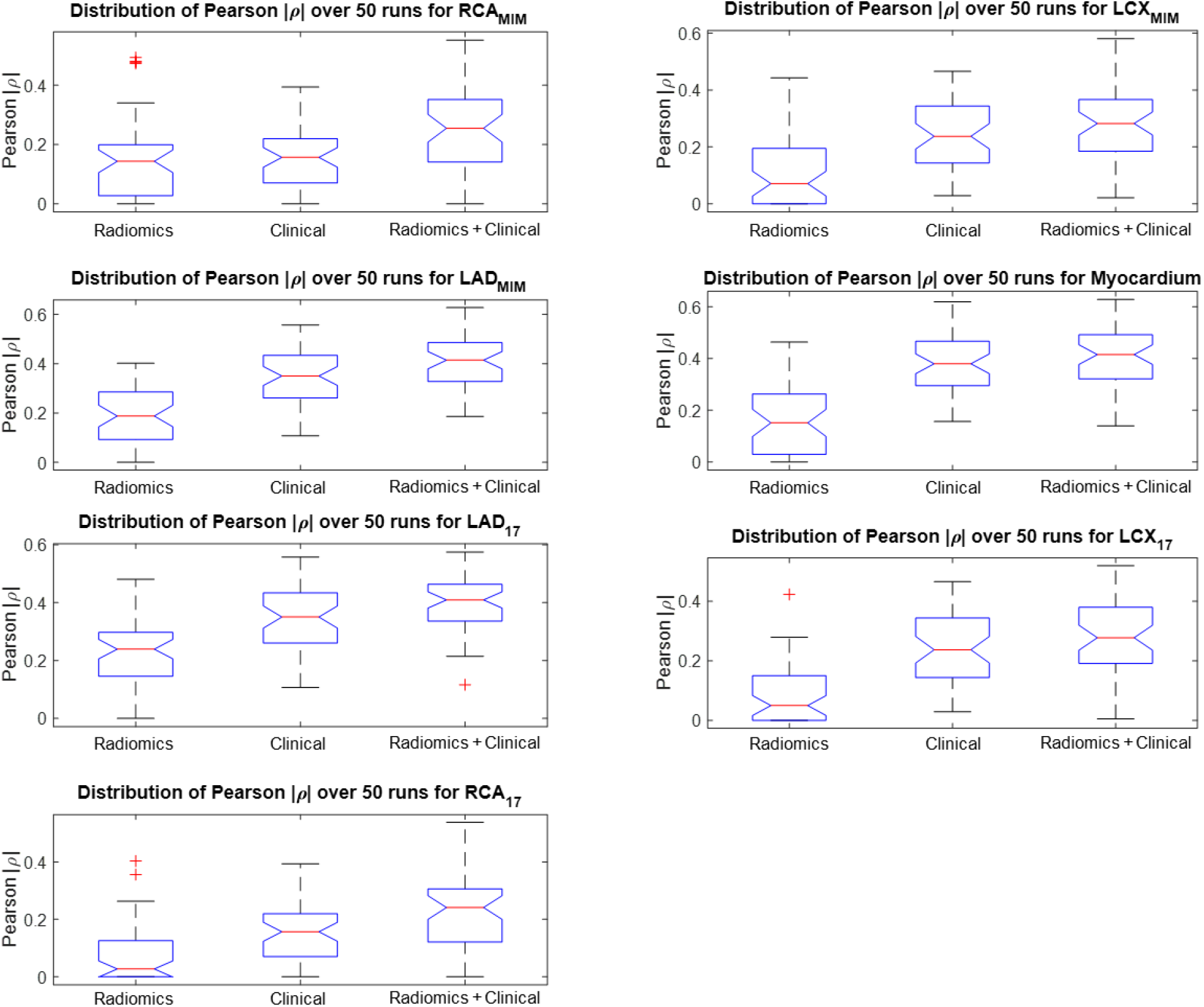
Distribution of absolute values of the Pearson’s *ρ* of the best fit out of 50 randomized trials of stepwise linear regression for radiomics, clinical and combined features, and for all 7 segmentations (the higher, the better). Adding radiomics to clinical features increases the correlation to the CAC score of the corresponding ROI.

**Figure 9.**
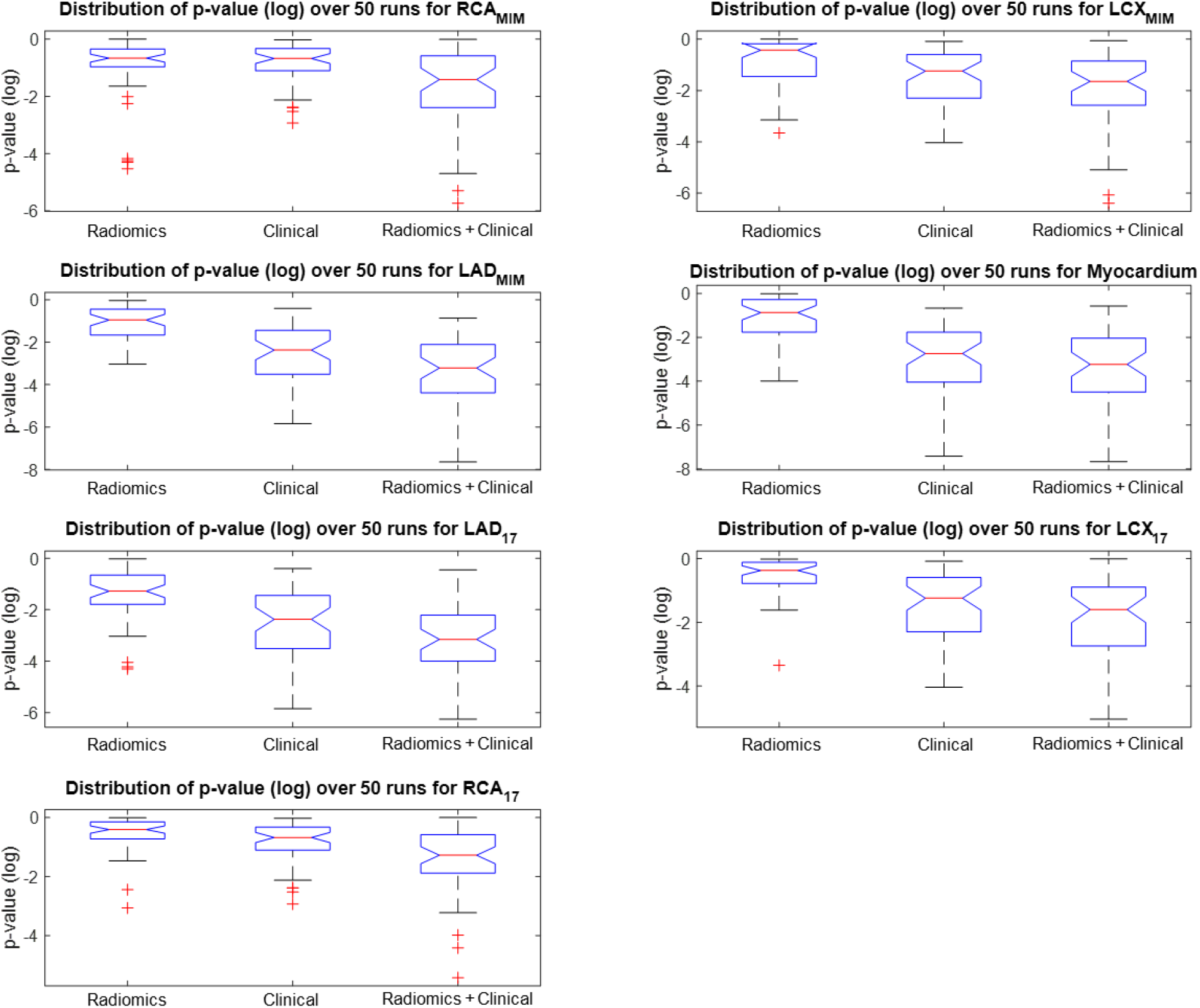
Distribution of p-values (log-scale) of the best fit out of 50 randomized trials of stepwise linear regression for radiomics, clinical and combined features, and for all 7 segmentations (the lower, the better). Adding radiomics to clinical features is seen to enhance the regression significance across all segmentations.

In clinical configurations, the most prevalent features in the best fit were somehow consistent across different segmentations and included age, hyperlipidemia, hypertension, and smoking. In the combined configuration, usually, the age variable was the first in the fit, followed by hyperlipidemia, GLSZM-small zone large GL emphasis, and GLDZM-short distance large GL emphasis.

## 4 Discussion

The current proposal is, to our knowledge, the first demonstration of employing radiomics of normal MPSS to predict CAC score as derived from a CT scan. In fact, to our knowledge, our team has been the first to study radiomics of cardiac SPECT imaging [50, 52, 53]. Our effort incorporates readily reconstructed 3D images and preserves the voxel intensities (instead of segmental polar map analyses). Recently, few studies have investigated the use of deep learning to predict CAD [54-56]; nonetheless, no studies, to our knowledge exist on predicting CAC scores from SPECT scans.

Our study fulfills a majority of “quality factors in radiomics studies”[24]: Vallières *et al*. have published a guideline for improving the quality of radiomics studies [24] in order to improve the reporting quality and the reproducibility of radiomics studies. The article includes a list of quality factors (Table 2 in [24]) proposed by the authors to help evaluate the quality of radiomics studies. A detailed explanation of how this study has implemented a majority of radiomics factors is explained in our Table S.3 (supplementary section **Error! Reference source not found**.). We add that a recent independent study comparing the performance of several IBSI-certified radiomics software packages [57]; SERA was shown to have the second-lowest difference between computed features and the IBSI benchmark values, further validating our SERA software package.

An interesting application of our proposed method is in cases of normal MPSS exams where a patient depicts significant CAC yet no pathology is detected from the MPSS image. Figure 10 shows such an example of an MPSS scan image in a polar plot format, i.e. 2D projection of the 3D SPECT image into its apex (center circle). This image was interpreted as normal by consensus, due to the absence of any reversibility and/or defect. At the same time, a diagnostic CT scan of this patient showed enormous calcification in the coronary arteries, representing an outstanding CAC score of 2239. Our multivariate analysis correctly stratified this patient’s CAC at the very high-risk level (Group 5, CAC>400 in Figure 4). This case ended up in 332 test sets with successful fits from 20×50 runs. The algorithm predicted an average CAC score of 1593 [639, 2037] across the 332 runs. Note that even the minimum computed CAC value (639) across 332 runs is still in group 5 of Figure 4 (CAC>400), flagging the patient with increased cardiovascular risk. Further investigation of this case demonstrates how the machine learning algorithm correctly predicted this elevated CAC score: the patient was positive for almost all clinical factors, including diabetes, hypertension, hyperlipidemia, family history of cardiac disease, with a soaring BMI of 31.5, in addition to heterogeneities that got captured by higher level radiomic features. In fact, the patient’s obesity might have made the impression that the slight visual heterogeneity had been an overcorrection artifact due to the patient’s obesity. Despite all that, the MPSS scan was read as normal by consensus, whereas the patient was associated with an increased cardiovascular risk due to enormous plaque burden that correspondingly required careful planning for his/her monitoring and treatment This is an interesting example that demonstrates the promise of our proposed research to assist in finding such cases with elevated CAC scores that otherwise might have failed to be noticed without the need to undergo a diagnostic CT scan.

**Figure 10.**
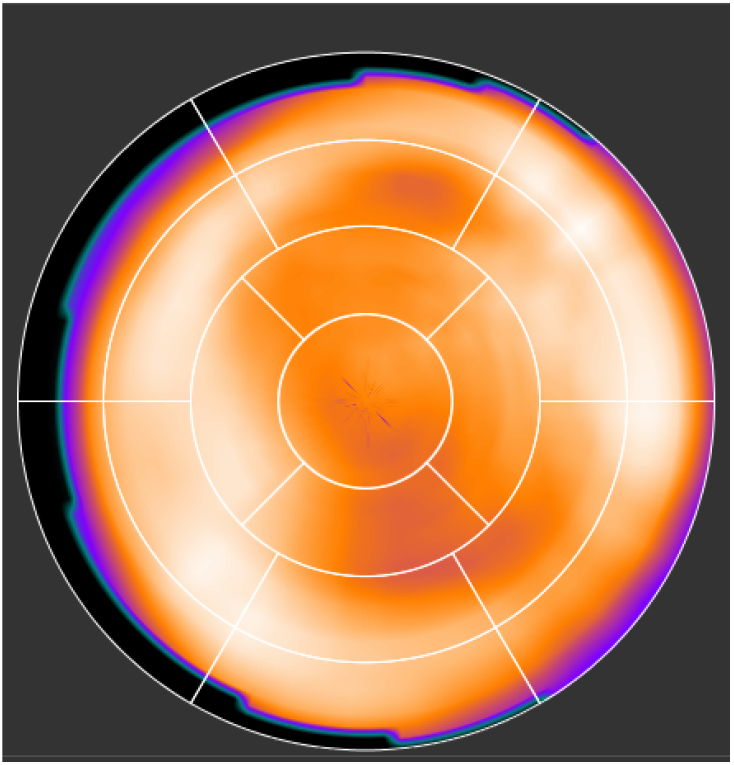
A normal MPSS with severe calcification. This scan was reported as normal by consensus, due to relatively uniform uptake with no reversibility and/or fixed defect; yet the diagnostic CAC CT scan showed an enormous CAC score of 2239. Our multivariate analysis predicted a CAC value with an average of 1593 [639, 2037] using combined radiomic and clinical features.

The study of MPSS radiomics is a challenging task due to several reasons. First, SPECT is a low-resolution imaging modality that results in a substantial loss of heterogeneity information that had the potential to provide extra knowledge about the blood flow and other functionalities of the heart that could have captured by radiomics. Moreover, the lack of quantitation in SPECT imaging further causes loss of information, resulting in a mostly-qualitative interpretation of the scan. The absence of quantitation prevents the utilization of several useful radiomic features, mainly, popular first-order intensity-based features. It is possible to perform quantitative SPECT imaging [58, 59] though this commonly requires dynamic imaging followed by kinetic modeling, resulting in added clinical value [60]. In any case, it is interesting to investigate whether radiomics coupled with standard SPECT, as routinely performed in the clinic, can unravel certain information that so far has only been yielded by quantitative SPECT. Furthermore, it can be hypothesized that quantitative SPECT coupled with radiomics can lead to further effective biomarkers of disease.

Another significant contributor to the challenges in SPECT radiomics is heterogeneity caused by inherent artifacts of SPECT imaging. MPSS, specifically, can cause artifacts on the reconstructed image that can appear as reduced uptake in the image, an example of which is shown in Figure 11. This effect, referred to as apical thinning, is a well-known phenomenon in MPS. It is often attributed to a reduced myocardial thickness at the apex of the left ventricle. Attenuation correction during the reconstruction appears to exaggerate this effect [61]. Moreover, soft tissue attenuation artifacts also impact MP SPECT images [62]. These artifacts generally appear as fixed defects. Attenuation due to breast tissue usually results in a perfusion defect along the anterior wall of the left ventricle, also affecting the lateral wall, septum, and apex [63]. The effect would be similar to that in Figure 11. During our data collection phase, we observed several cases with this effect apparent in their reconstructed image, and mild-to-severe ones were excluded by our physician. The heterogeneity caused by this effect may be captured by the radiomic features, while it is irrelevant to calcifications in arteries (a confound), and our multivariate scheme is unable to distinguish and exclude such cases. More advanced multivariate schemes such as deep neural networks trained with very large datasets might be able to accurately categorize these examples. That is why any clinically related decisions and results deduced from radiomics and machine learning analyses such as ours is only complimentary to clinical decision-making procedure and should be ultimately verified by an experienced physician.

**Figure 11.**
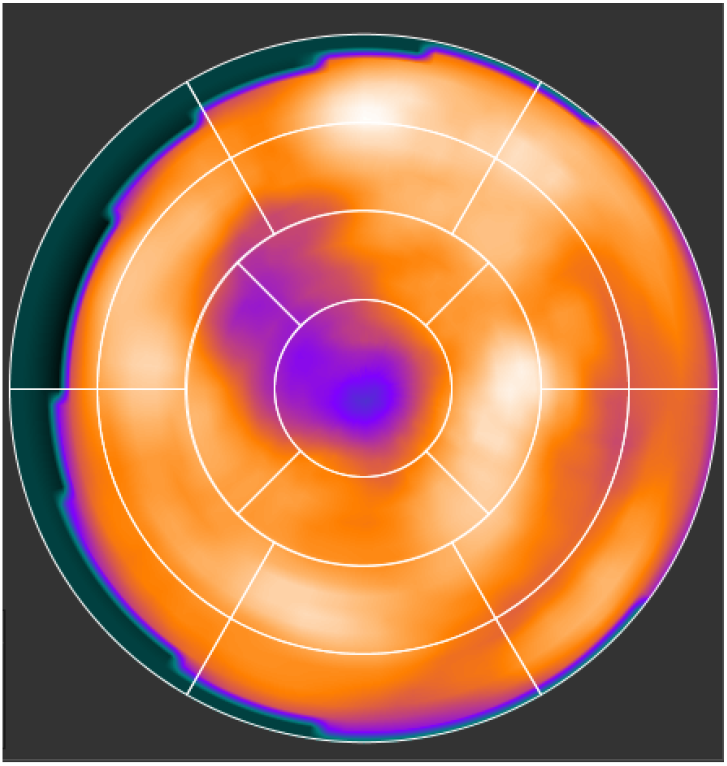
A normal MPSS with apical thinning.

Our results interestingly demonstrated biological correlate with radiomic features. Two features frequently survived regression training/validation, ending up in the multivariate fit, namely GLSZM-small zone high GL emphasis and GLDZM-short distance high GL emphasis. Both features emphasize higher GLs, i.e., more weight goes to higher voxel values, and in MP SPECT images, higher GLs in a discretized SPECT image depict higher blood flow. This is interesting to observe and seems intuitive that radiomic features that may capture higher blood flow and are more sensitive to high blood flow variations in a small region surrounding each voxel in each cardiac segment, are selected. Hyperlipidemia and age were the two most frequent clinical features appearing in the fit. This also has an intuition associated with it, since the amount and prevalence of CAC reportedly increase steadily with age [64]. Also, hyperlipidemia is shown to be associated with high CAC score as well as multivessel CAD independent of cardiac risk factors [65].

In the current study, even after feature elimination steps (which resulted in considerable reductions by a factor of 70), univariate analysis following correction for false discovery was not able to discover significant correlations with outcome. By contrast, our multivariate analysis (designed to mitigate overfitting) was able to make significant prediction in all segments of the heart. Our results demonstrated the possibility of predicting the CAC score significantly utilizing a standardized and reproducible framework from a combined set of radiomics and clinical features. Future studies incorporating more advanced multivariate analyses including deep neural networks can be pursued to potentially enhance predictive power for CAC scores. In this work, we started with an image dataset acquired from scanners from a diverse set of vendors, subsequently calculating standardized radiomic features incorporating a diverse set of grey levels, ultimately deducing a subset of 56 robust features with one robust grey level “independent of outcome”. These intermediate study outcomes can also be adopted by future researchers and contribute to future more general radiomics studies of MP SPECT images.

## 5 Conclusion

This study investigates the hypothesis that radiomics analysis of MPSS images may convey information regarding calcification in coronary arteries. This can be especially significant as CAC is not commonly reimbursed (e.g. by the CMS) nor widely deployed in community settings. We segmented MPSS images into LAD, RCA, LCX, each with two varieties, as well as the entire myocardium, evaluating features for all 7 segments, employing our standardized SERA radiomics package. Our dataset consisted of 428 patients with normal (non-ischemic) MPSS images verified to be free from artifact or spillover, in addition to their detailed CAC score acquired from CT, and other clinical demographics. Our focus was on patients with normal stress scan given the significant potential clinical value for prediction of coronary artery calcification from such images. Through a multi-step blind-to-outcome unsupervised feature selection phase, we significantly reduced our feature space ∼70 folds. Our univariate analysis between each feature and the corresponding CAC score did not demonstrate significant results. Our multivariate analysis, however, was able to significantly predict the CAC score of all cardiac segments when combining radiomic features with clinical features. Our method has the potential to identify cases with high coronary artery calcification from normal MPSS images, that can be prompted for more appropriate care, suggesting that radiomics analysis adds diagnostic and prognostic value to standard MP SPECT for wide clinical usage.

## Data Availability

The radiomics framework used for this study, standardized environment of radiomics anlaysis (SEAR) is available via https://github.com/ashrafinia/SERA.
The Matlab code of multivariate analysis will be included in the submitted version.

## Code availability

Our SERA software, developed in the course of this effort, is publicly available at https://github.com/ashrafinia/SERA.

## Acknowledgement

This work was in part supported by a Bradley-Alavi fellowship award to the first author from the Society of Nuclear Medicine & Molecular Imaging (SNMMI). The authors gratefully acknowledge very helpful support from colleagues at MIM Software Inc. The authors also would like to thank Dr. Payam Ghazi MD, Dr. Charles Marcus MD, Dr. Mehdi Taghipour MD, Dr. Rongkai Yen MD, and Dr. Martin Lodge Ph.D. for their support during this study.

