## Supplementary for "Radiomics Analysis of Clinical Myocardial Perfusion Stress SPECT Images to Identify Coronary Artery Calcification"

### Supplementary Materials

#### S.1. Additional statistics on patients in this study

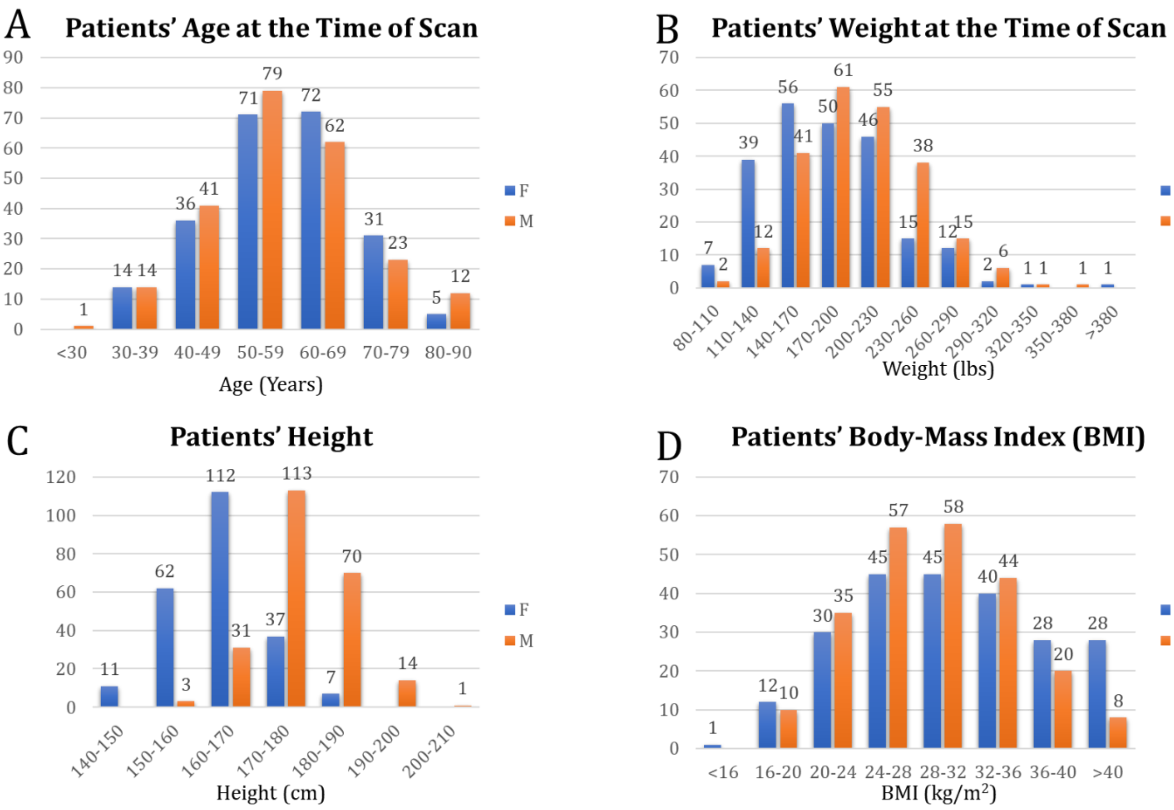


*Figure S.1. Distribution of A) age, B) weight, C) height and D) BMI at the time of scan grouped into male (orange) and female (blue)*

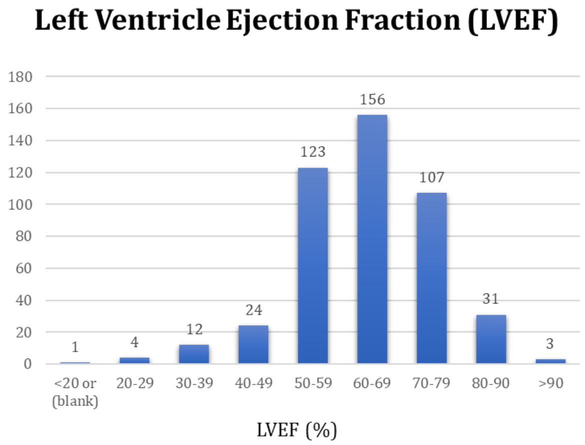


*Figure S.2. Distribution of LVEF in patients of the dataset*

#### S.2. Standardized Environment for Radiomics Analysis (SERA)

##### S.2.1 Introduction

The Standardized Environment for Radiomics Analysis (SERA) is a Matlab®-based framework developed at Johns Hopkins University that calculates radiomic features based on guidelines from the Image Biomarker Standardization Initiative (IBSI) (https://arxiv.org/pdf/1612.07003.pdf). SERA is capable of processing images from various clinical imaging modalities such as CT, MRI, PET, and SPECT. Radiomic features calculated with SERA are standardized and in compliance with IBSI, which ensures their reproducibility.

##### S.2.2 Radiomic Features

SERA calculates 487 IBSI-standardized features (as outlined in Table below). These include 79 first-order features (morphology, statistical, histogram, and intensity-histogram features), 272 higher-order 2D features, and 136 3D features. Different subsets of features can be selected, such as the default of 215 features (first order + higher-order 3D).

*Table S.1. Radiomic features SERA calculates in each feature family. Different 2D, 2.5D, and 3D configurations are explained in detail in the IBSI guideline. Users can set to return only a selected subset of these features.*

| Feature Family | Subtypes | Number of Features |
|---|---|---|
| Morphology | - | 29 |
| Local Intensity | - | 2 |
| Intensity-based Statistics | - | 18 |
| Intensity Histogram | - | 23 |
| Intensity-Volume Histogram | - | 7 |
| Gray Level Co-occurrence Matrix (GLCM) | 2D Averaged<br>2D Slice-Merged<br>2.5D Direction Merged<br>2.5 D All Merged<br>3D Averaged<br>3D Merged | 25<br>25<br>25<br>25<br>25<br>25 |
| Gray Level Run Length Matrix (GLRLM) | 2D Averaged<br>2D Slice-Merged<br>2.5D Direction Merged<br>2.5 D All Merged<br>3D Averaged<br>3D Merged | 16<br>16<br>16<br>16<br>16<br>16 |
| Gray Level Size Zone Matrix (GLZSM) | 2D<br>2.5 D<br>3D | 16<br>16<br>16 |
| Gray Level Distance Zone Matrix (GLDZM) | 2D<br>2.5 D<br>3D | 16<br>16<br>16 |
| Neighborhood Grey Tone Difference Matrix (NGTDM) | 2D<br>2.5 D<br>3D | 5<br>5<br>5 |
| Neighboring Grey Level Dependence Matrix (NGLDM) | 2D<br>2.5 D<br>3D | 17<br>17<br>17 |
| **Total** | | **487** |

##### S.2.3 Feature Evaluation Settings

SERA has options to set and modify all parameters defined or used in the IBSI guideline. The following parameters can be set up in the image preparation setting of SERA (for detailed information please refer to IBSI documentation [1]):

- Resampling and interpolation:
  - resample to 2D and 3D isotropic voxel sizes; interpolation algorithm used in resampling image and ROI (nearest/linear/cubic); partial volume threshold (mostly used for CT HU).
- Discretization:
  - bin size, discretization type (fixed bin size/fixed bin numbers), discretization algorithm (uniform/Lloyd)
- Other:
  - grey-level rounding, image re-segmentation (range re-segmentation, outliers' re-segmentation)

##### S.2.4 Radiomic feature values for IBSI benchmark datasets as calculated by SERA

IBSI shared two phantoms between all participating institutions in two phases ROI to facilitate the process of establishing reference values for features. In phase I, it was a small 80-voxel three-dimensional digital phantom with a 74-voxel ROI mask to facilitate the process of establishing reference values for features, without involving image processing. In phase II, a publicly available CT image in a patient with lung cancer with an accompanying gross tumor volume as the ROI [1, 2]. We have included a supplemental spreadsheet containing the features calculated by SERA on the IBSI benchmark CT phantom in the supplementary materials. Table S.2 contains the values of each feature computed by SERA in comparison to the IBSI reported benchmark values for Configuration D of the CT phantom as indicated in IBSI guideline [1], which is the closest configuration to the setup in the current study. The results in this spreadsheet demonstrate the compliance of SERA with IBSI radiomics guidelines.

*Table S.2. List of radiomics features calculated by SERA for Configuration D of IBSI CT phantom [1]. SERA shows great compliance with IBSI with 99.8% match in calculated results.*

| Family | Image_Biomarker | Benchmark Value | Tolerance | SERA Result | Diff | Check |
|---|---|---|---|---|---|---|
| Morphology | Volume (mesh-based) | 367000 | 6000 | 367453.66 | 0 | match |
| Morphology | Volume (counting) | 368000 | 6000 | 367880 | 0 | match |
| Morphology | Surface area | 34300 | 400 | 34306.254 | 0 | match |
| Morphology | Surface to volume ratio | 0.0934 | 0.0007 | 0.093 | 0 | match |
| Morphology | Compactness 1 | 0.0326 | 0.0002 | 0.033 | 0 | match |
| Morphology | Compactness 2 | 0.378 | 0.004 | 0.378 | 0 | match |
| Morphology | Spherical disproportion | 1.38 | 0.01 | 1.383 | 0 | match |
| Morphology | Sphericity | 0.723 | 0.003 | 0.723 | 0 | match |
| Morphology | Asphericity | 0.383 | 0.004 | 0.383 | 0 | match |
| Morphology | Centre of mass shift | 64.9 | 2.8 | 64.926 | 0 | match |
| Morphology | Maximum 3D diameter | 125 | 1 | 125.06 | 0 | match |
| Morphology | Major axis length | 93.3 | 0.5 | 93.27 | 0 | match |
| Morphology | Minor axis length | 82 | 0.5 | 82.005 | 0 | match |
| Morphology | Least axis length | 70.9 | 0.4 | 70.902 | 0 | match |
| Morphology | Elongation | 0.879 | 0.001 | 0.879 | 0 | match |
| Morphology | Flatness | 0.76 | 0.001 | 0.76 | 0 | match |

| | | | | | | |
|---|---|---|---|---|---|---|
| Morphology | Volume density (AABB) | 0.478 | 0.003 | 0.478 | 0 | match |
| Morphology | Area density (AABB) | 0.678 | 0.003 | 0.678 | 0 | match |
| Morphology | Volume density (OMBB) | N. A. | N. A. | 0.526 | | N. A. |
| Morphology | Area density (OMBB) | N. A. | N. A. | 0.723 | | N. A. |
| Morphology | Volume density (AEE) | 1.29 | 0.01 | 1.294 | 0 | match |
| Morphology | Area density (AEE) | 1.62 | 0.01 | 1.605 | 0.01 | match |
| Morphology | Volume density (MVEE) | N. A. | N. A. | 0.615 | | N. A. |
| Morphology | Area density (MVEE) | N. A. | N. A. | 1.121 | | N. A. |
| Morphology | Volume density (convex hull) | 0.834 | 0.002 | 0.834 | 0 | match |
| Morphology | Area density (convex hull) | 1.13 | 0.01 | 1.13 | 0 | match |
| Morphology | Integrated intensity | -8640000 | 1560000 | -8641752 | 0 | match |
| Morphology | Moran's I index | 0.0622 | 0.0013 | 0.062 | 0 | match |
| Morphology | Geary's C measure | 0.851 | 0.001 | 0.851 | 0 | match |
| Local intensity | Local intensity peak | 201 | 10 | 200.821 | 0 | match |
| Local intensity | Global intensity peak | N. A. | N. A. | 200.821 | | N. A. |
| Statistics | Mean | -23.5 | 3.9 | -23.518 | 0 | match |
| Statistics | Variance | 32800 | 2100 | 32786.871 | 0 | match |
| Statistics | Skewness | -2.28 | 0.06 | -2.28 | 0 | match |
| Statistics | (Excess) kurtosis | 4.35 | 0.32 | 4.351 | 0 | match |
| Statistics | Median | 42 | 0.4 | 42 | 0 | match |
| Statistics | Minimum | -724 | 12 | -724 | 0 | match |
| Statistics | 10th percentile | -304 | 20 | -304 | 0 | match |
| Statistics | 90th percentile | 86 | 0.1 | 86 | 0 | match |
| Statistics | Maximum | 521 | 22 | 521 | 0 | match |
| Statistics | Interquartile range | 57 | 4.1 | 57 | 0 | match |
| Statistics | Range | 1240 | 40 | 1245 | 0 | match |
| Statistics | Mean absolute deviation | 123 | 6 | 122.543 | 0 | match |
| Statistics | Robust mean absolute deviation | 46.8 | 3.6 | 46.827 | 0 | match |
| Statistics | Median absolute deviation | 94.7 | 3.8 | 94.73 | 0 | match |
| Statistics | Coefficient of variation | -7.7 | 1.01 | -7.699 | 0 | match |
| Statistics | Quartile coefficient of dispersion | 0.74 | 0.011 | 0.74 | 0 | match |
| Statistics | Energy | 1.48e9 | 1.4e9 | 1.48e9 | 0 | match |
| Statistics | Root mean square | 183 | 7 | 182.592 | 0 | match |
| Intensity histogram | Mean | 18.5 | 0.5 | 18.503 | 0 | match |
| Intensity histogram | Variance | 21.7 | 0.4 | 21.69 | 0 | match |
| Intensity histogram | Skewness | -2.27 | 0.06 | -2.268 | 0 | match |
| Intensity histogram | Kurtosis | 4.31 | 0.32 | 4.308 | 0 | match |
| Intensity histogram | Median | 20 | 0.5 | 20 | 0 | match |
| Intensity histogram | Minimum | 1 | 0 | 1 | 0 | match |
| Intensity histogram | 10th percentile | 11 | 0.7 | 11 | 0 | match |
| Intensity histogram | 90th percentile | 21 | 0.5 | 21 | 0 | match |
| Intensity histogram | Maximum | 32 | 0 | 32 | 0 | match |
| Intensity histogram | Mode | 20 | 0.4 | 20 | 0 | match |
| Intensity histogram | Interquartile range | 2 | 0.06 | 2 | 0 | match |
| Intensity histogram | Range | 31 | 0 | 31 | 0 | match |
| Intensity histogram | Mean absolute deviation | 3.15 | 0.05 | 3.151 | 0 | match |
| Intensity histogram | Robust mean absolute deviation | 1.33 | 0.06 | 1.328 | 0 | match |
| Intensity histogram | Median absolute deviation | 2.41 | 0.04 | 2.407 | 0 | match |
| Intensity histogram | Coefficient of variation | 0.252 | 0.006 | 0.252 | 0 | match |
| Intensity histogram | Quartile coefficient of dispersion | 0.05 | 0.0021 | 0.05 | 0 | match |
| Intensity histogram | Entropy | 2.94 | 0.01 | 2.94 | 0 | match |
| Intensity histogram | Uniformity | 0.229 | 0.003 | 0.229 | 0 | match |
| Intensity histogram | Maximum histogram gradient | 7260 | 200 | 7263 | 0 | match |
| Intensity histogram | Maximum gradient grey level | 19 | 0.4 | 19 | 0 | match |
| Intensity histogram | Minimum histogram gradient | -6670 | 230 | -6674 | 0 | match |
| Intensity histogram | Minimum gradient grey level | 22 | 0.4 | 22 | 0 | match |
| Intensity vol histogram | Volume fraction at 10% intensity | 0.972 | 0.003 | 0.972 | 0 | match |
| Intensity vol histogram | Volume fraction at 90% intensity | 0.00009 | 0.000415 | 0 | 0 | match |
| Intensity vol histogram | Intensity at 10% volume | 87 | 0.1 | 87 | 0 | match |
| Intensity vol histogram | Intensity at 90% volume | -303 | 20 | -303 | 0 | match |
| Intensity vol histogram | Volume fraction diff between 10% and 90% intensity | 0.971 | 0.001 | 0.971 | 0 | match |
| Intensity vol histogram | Intensity difference between 10% and 90% volume | 390 | 20 | 390 | 0 | match |
| Intensity vol histogram | Area under the IVH curve | 0.563 | 0.012 | 0.563 | 0 | match |

| | | | | | | |
|---|---|---|---|---|---|---|
| GLCM (3D, averaged) | Joint maximum | 0.232 | 0.007 | 0.232 | 0 | match |
| GLCM (3D, averaged) | Joint average | 18.9 | 0.5 | 18.852 | 0 | match |
| GLCM (3D, averaged) | Joint variance | 17.6 | 0.4 | 17.628 | 0 | match |
| GLCM (3D, averaged) | Joint entropy | 4.95 | 0.03 | 4.947 | 0 | match |
| GLCM (3D, averaged) | Difference average | 1.29 | 0.01 | 1.293 | 0 | match |
| GLCM (3D, averaged) | Difference variance | 5.37 | 0.11 | 5.369 | 0 | match |
| GLCM (3D, averaged) | Difference entropy | 2.13 | 0.01 | 2.134 | 0 | match |
| GLCM (3D, averaged) | Sum average | 37.7 | 0.8 | 37.705 | 0 | match |
| GLCM (3D, averaged) | Sum variance | 63.4 | 1.3 | 63.441 | 0 | match |
| GLCM (3D, averaged) | Sum entropy | 3.68 | 0.02 | 3.676 | 0 | match |
| GLCM (3D, averaged) | Angular second moment | 0.11 | 0.003 | 0.11 | 0 | match |
| GLCM (3D, averaged) | Contrast | 7.07 | 0.13 | 7.071 | 0 | match |
| GLCM (3D, averaged) | Dissimilarity | 1.29 | 0.01 | 1.293 | 0 | match |
| GLCM (3D, averaged) | Inverse difference | 0.682 | 0.003 | 0.682 | 0 | match |
| GLCM (3D, averaged) | Inverse difference normalized | 0.965 | 0.001 | 0.965 | 0 | match |
| GLCM (3D, averaged) | Inverse difference moment | 0.656 | 0.003 | 0.656 | 0 | match |
| GLCM (3D, averaged) | Inverse difference moment normalized | 0.994 | 0.001 | 0.994 | 0 | match |
| GLCM (3D, averaged) | Inverse variance | 0.341 | 0.005 | 0.341 | 0 | match |
| GLCM (3D, averaged) | Correlation | 0.798 | 0.005 | 0.798 | 0 | match |
| GLCM (3D, averaged) | Autocorrelation | 370 | 16 | 369.511 | 0 | match |
| GLCM (3D, averaged) | Cluster tendency | 63.4 | 1.3 | 63.441 | 0 | match |
| GLCM (3D, averaged) | Cluster shade | -1270 | 40 | -1272.93 | 0 | match |
| GLCM (3D, averaged) | Cluster prominence | 35700 | 1400 | 35664.719 | 0 | match |
| GLCM (3D, averaged) | Information correlation 1 | -0.231 | 0.003 | -0.231 | 0 | match |
| GLCM (3D, averaged) | Information correlation 2 | 0.845 | 0.003 | 0.845 | 0 | match |
| GLCM (3D, merged) | Joint maximum | 0.232 | 0.007 | 0.232 | 0 | match |
| GLCM (3D, merged) | Joint average | 18.9 | 0.5 | 18.852 | 0 | match |
| GLCM (3D, merged) | Joint variance | 17.6 | 0.4 | 17.638 | 0 | match |
| GLCM (3D, merged) | Joint entropy | 4.96 | 0.03 | 4.965 | 0 | match |
| GLCM (3D, merged) | Difference average | 1.29 | 0.01 | 1.29 | 0 | match |
| GLCM (3D, merged) | Difference variance | 5.38 | 0.11 | 5.381 | 0 | match |
| GLCM (3D, merged) | Difference entropy | 2.14 | 0.01 | 2.139 | 0 | match |
| GLCM (3D, merged) | Sum average | 37.7 | 0.8 | 37.703 | 0 | match |
| GLCM (3D, merged) | Sum variance | 63.5 | 1.3 | 63.506 | 0 | match |
| GLCM (3D, merged) | Sum entropy | 3.68 | 0.02 | 3.679 | 0 | match |
| GLCM (3D, merged) | Angular second moment | 0.109 | 0.003 | 0.109 | 0 | match |
| GLCM (3D, merged) | Contrast | 7.05 | 0.13 | 7.045 | 0 | match |
| GLCM (3D, merged) | Dissimilarity | 1.29 | 0.01 | 1.29 | 0 | match |
| GLCM (3D, merged) | Inverse difference | 0.682 | 0.003 | 0.682 | 0 | match |
| GLCM (3D, merged) | Inverse difference normalized | 0.965 | 0.001 | 0.965 | 0 | match |
| GLCM (3D, merged) | Inverse difference moment | 0.657 | 0.003 | 0.657 | 0 | match |
| GLCM (3D, merged) | Inverse difference moment normalized | 0.994 | 0.001 | 0.994 | 0 | match |
| GLCM (3D, merged) | Inverse variance | 0.34 | 0.005 | 0.34 | 0 | match |
| GLCM (3D, merged) | Correlation | 0.8 | 0.005 | 0.8 | 0 | match |
| GLCM (3D, merged) | Autocorrelation | 370 | 16 | 369.503 | 0 | match |
| GLCM (3D, merged) | Cluster tendency | 63.5 | 1.3 | 63.506 | 0 | match |
| GLCM (3D, merged) | Cluster shade | -1280 | 40 | -1275.262 | 0 | match |
| GLCM (3D, merged) | Cluster prominence | 35700 | 1500 | 35742.844 | 0 | match |
| GLCM (3D, merged) | Information correlation 1 | -0.225 | 0.003 | -0.225 | 0 | match |
| GLCM (3D, merged) | Information correlation 2 | 0.846 | 0.003 | 0.846 | 0 | match |
| GLRLM (3D, averaged) | Short runs emphasis | 0.734 | 0.001 | 0.734 | 0 | match |
| GLRLM (3D, averaged) | Long runs emphasis | 6.66 | 0.18 | 6.657 | 0 | match |
| GLRLM (3D, averaged) | Low GL run emphasis | 0.0257 | 0.0012 | 0.026 | 0 | match |
| GLRLM (3D, averaged) | High GL run emphasis | 326 | 17 | 325.741 | 0 | match |
| GLRLM (3D, averaged) | Short run low GL emphasis | 0.0232 | 0.001 | 0.023 | 0 | match |
| GLRLM (3D, averaged) | Short run high GL emphasis | 219 | 13 | 218.622 | 0 | match |
| GLRLM (3D, averaged) | Long run low GL emphasis | 0.0484 | 0.0031 | 0.048 | 0 | match |
| GLRLM (3D, averaged) | Long run high GL emphasis | 2670 | 30 | 2667.075 | 0 | match |
| GLRLM (3D, averaged) | GL non-uniformity | 3290 | 10 | 3293.649 | 0 | match |
| GLRLM (3D, averaged) | GL non-uniformity normalized | 0.133 | 0.002 | 0.133 | 0 | match |
| GLRLM (3D, averaged) | Run length non-uniformity | 12400 | 200 | 12351.102 | 0 | match |
| GLRLM (3D, averaged) | Run length non-uniformity normalized | 0.5 | 0.001 | 0.5 | 0 | match |
| GLRLM (3D, averaged) | Run percentage | 0.554 | 0.005 | 0.554 | 0 | match |

| | | | | | | |
|---|---|---|---|---|---|---|
| GLRLM (3D, averaged) | GL variance | 31.5 | 0.4 | 31.453 | 0 | match |
| GLRLM (3D, averaged) | Run length variance | 3.35 | 0.14 | 3.348 | 0 | match |
| GLRLM (3D, averaged) | Run entropy | 5.08 | 0.02 | 5.081 | 0 | match |
| GLRLM (3D, merged) | Short runs emphasis | 0.736 | 0.001 | 0.736 | 0 | match |
| GLRLM (3D, merged) | Long runs emphasis | 6.56 | 0.18 | 6.556 | 0 | match |
| GLRLM (3D, merged) | Low GL run emphasis | 0.0257 | 0.0012 | 0.026 | 0 | match |
| GLRLM (3D, merged) | High GL run emphasis | 326 | 17 | 326.073 | 0 | match |
| GLRLM (3D, merged) | Short run low GL emphasis | 0.0232 | 0.001 | 0.023 | 0 | match |
| GLRLM (3D, merged) | Short run high GL emphasis | 219 | 13 | 219.402 | 0 | match |
| GLRLM (3D, merged) | Long run low GL emphasis | 0.0478 | 0.0031 | 0.048 | 0 | match |
| GLRLM (3D, merged) | Long run high GL emphasis | 2630 | 30 | 2625.593 | 0 | match |
| GLRLM (3D, merged) | GL non-uniformity | 42800 | 200 | 42767.969 | 0 | match |
| GLRLM (3D, merged) | GL non-uniformity normalized | 0.134 | 0.002 | 0.134 | 0 | match |
| GLRLM (3D, merged) | Run length non-uniformity | 160000 | 3000 | 160418.5 | 0 | match |
| GLRLM (3D, merged) | Run length non-uniformity normalized | 0.501 | 0.001 | 0.501 | 0 | match |
| GLRLM (3D, merged) | Run percentage | 0.554 | 0.005 | 7.199 | 6.66 | no match |
| GLRLM (3D, merged) | GL variance | 31.4 | 0.4 | 31.425 | 0 | match |
| GLRLM (3D, merged) | Run length variance | 3.29 | 0.13 | 3.295 | 0 | match |
| GLRLM (3D, merged) | Run entropy | 5.08 | 0.02 | 5.083 | 0 | match |
| GLZSM(3D) | Small zone emphasis | 0.637 | 0.005 | 0.637 | 0 | match |
| GLZSM(3D) | Large zone emphasis | 99100 | 2800 | 99078.516 | 0 | match |
| GLZSM(3D) | Low GL emphasis | 0.0409 | 0.0005 | 0.041 | 0 | match |
| GLZSM(3D) | High GL emphasis | 188 | 10 | 188.183 | 0 | match |
| GLZSM(3D) | Small zone low GL emphasis | 0.0248 | 0.0004 | 0.025 | 0 | match |
| GLZSM(3D) | Small zone high GL emphasis | 117 | 7 | 116.553 | 0 | match |
| GLZSM(3D) | Large zone low GL emphasis | 241 | 14 | 240.778 | 0 | match |
| GLZSM(3D) | Large zone high GL emphasis | 41400000 | 300000 | 41404348 | 0 | match |
| GLZSM(3D) | GL non-uniformity | 212 | 6 | 212.134 | 0 | match |
| GLZSM(3D) | GL non uniformity normalized | 0.0491 | 0.0008 | 0.049 | 0 | match |
| GLZSM(3D) | Zone size non-uniformity | 1630 | 10 | 1629.113 | 0 | match |
| GLZSM(3D) | Zone size non-uniformity normalized | 0.377 | 0.006 | 0.377 | 0 | match |
| GLZSM(3D) | Zone percentage | 0.0972 | 0.0007 | 0.097 | 0 | match |
| GLZSM(3D) | GL variance | 32.7 | 1.6 | 32.718 | 0 | match |
| GLZSM(3D) | Zone size variance | 99000 | 2800 | 98972.773 | 0 | match |
| GLZSM(3D) | Zone size entropy | 6.52 | 0.01 | 6.515 | 0 | match |
| GLDZM(3D) | Small distance emphasis | 0.579 | 0.004 | 0.579 | 0 | match |
| GLDZM(3D) | Large distance emphasis | 10.3 | 0.1 | 10.258 | 0 | match |
| GLDZM(3D) | Low GL emphasis | 0.0409 | 0.0005 | 0.041 | 0 | match |
| GLDZM(3D) | High GL emphasis | 188 | 10 | 188.183 | 0 | match |
| GLDZM(3D) | Small distance low GL emphasis | 0.0302 | 0.0006 | 0.03 | 0 | match |
| GLDZM(3D) | Small distance high GL emphasis | 99.3 | 5.1 | 99.3 | 0 | match |
| GLDZM(3D) | Large distance low GL emphasis | 0.183 | 0.004 | 0.183 | 0 | match |
| GLDZM(3D) | Large distance high GL emphasis | 2620 | 110 | 2619.168 | 0 | match |
| GLDZM(3D) | GL non-uniformity | 212 | 6 | 212.134 | 0 | match |
| GLDZM(3D) | GL non-uniformity normalized | 0.0491 | 0.0008 | 0.049 | 0 | match |
| GLDZM(3D) | Zone distance non-uniformity | 1370 | 20 | 1369.445 | 0 | match |
| GLDZM(3D) | Zone distance non-uniformity normalized | 0.317 | 0.004 | 0.317 | 0 | match |
| GLDZM(3D) | Zone percentage | 0.0972 | 0.0007 | 0.097 | 0 | match |
| GLDZM(3D) | GL variance | 32.7 | 1.6 | 32.718 | 0 | match |
| GLDZM(3D) | Zone distance variance | 4.61 | 0.04 | 4.614 | 0 | match |
| GLDZM(3D) | Zone distance entropy | 6.61 | 0.03 | 6.614 | 0 | match |
| NGTDM (3D) | Coarseness | 0.000208 | 0.000004 | 0 | 0 | match |
| NGTDM (3D) | Contrast | 0.046 | 0.0005 | 0.046 | 0 | match |
| NGTDM (3D) | Busyness | 5.14 | 0.14 | 5.144 | 0 | match |
| NGTDM (3D) | Complexity | 400 | 5 | 399.694 | 0 | match |
| NGTDM (3D) | Strength | 0.162 | 0.008 | 0.162 | 0 | match |
| NGLDM(3D) | Low dependence emphasis | 0.0912 | 0.0007 | 0.091 | 0 | match |
| NGLDM(3D) | High dependence emphasis | 223 | 5 | 222.748 | 0 | match |
| NGLDM(3D) | Low GL count emphasis | 0.0168 | 0.0009 | 0.017 | 0 | match |
| NGLDM(3D) | High GL count emphasis | 364 | 16 | 364.049 | 0 | match |
| NGLDM(3D) | Low dependence low GL emphasis | 0.00357 | 0.00004 | 0.004 | 0 | match |
| NGLDM(3D) | Low dependence high GL emphasis | 18.9 | 1.1 | 18.945 | 0 | match |

| | | | | | | |
|---|---|---|---|---|---|---|
| NGLDM(3D) | High dependence low GL emphasis | 0.798 | 0.072 | 0.798 | 0 | match |
| NGLDM(3D) | High dependence high GL emphasis | 92800 | 1300 | 92761.625 | 0 | match |
| NGLDM(3D) | GL non-uniformity | 10200 | 300 | 10172.049 | 0 | match |
| NGLDM(3D) | GL non-uniformity normalized | 0.229 | 0.003 | 0.229 | 0 | match |
| NGLDM(3D) | Dependence count non-uniformity | 1840 | 30 | 1836.865 | 0 | match |
| NGLDM(3D) | Dependence count non-uniformity normalized | 0.0413 | 0.0003 | 0.041 | 0 | match |
| NGLDM(3D) | Dependence count percentage | 1 | 0 | 1 | 0 | match |
| NGLDM(3D) | GL variance | 21.7 | 0.4 | 21.69 | 0 | match |
| NGLDM(3D) | Dependence count variance | 63.9 | 1.3 | 63.923 | 0 | match |
| NGLDM(3D) | Dependence count entropy | 6.98 | 0.01 | 6.981 | 0 | match |
| NGLDM(3D) | Dependence count energy | 0.0113 | 0.0002 | 0.011 | 0 | match |

#### S.3. Matlab code of the multivariate stepwise regression outcome prediction analysis

We used the following code in Matlab 2019a for the procedure explained in section **Error! Reference source not found.**:Statistical Analysis: Outcome Prediction. It follows the calculation of the AIC criterion and selecting the best along with chi-squared (Fisher's method) to determine significance.

```
%%=========================================================================
%% Multivariate Stepwise Regression with Independent Test with p-value adjustment
%%=========================================================================
```

#### S.4. Radiomics Quality Factors

[3, 4]Table S.3 contains a cross-check list of radiomics quality factors, including details on how our study has implemented every factor [3].

*Table S.3. Radiomics quality factor implemented in the current study*

| | **Radiomics Quality Factor** | **Considered** | **Comment** |
|---|---|---|---|
| **Imaging** | Standardized imaging protocols | ✓ | A standardized protocol was in place across all Johns Hopkins Hospital SPECT scanners |
| | Imaging quality insurance | ✓ | Scanners were well-maintained and calibrated to produce high-quality clinical images |
| | Calibration | ✓ | Image post-processing and standardized radiomics calculation with SERA |
| **Experimental setup** | Multi-institutional /external datasets | ✗ | Left for future studies… |
| | Prospective study | ✗ | Left for future studies… |
| **Feature Selection** | Feature robustness | ✓ | Feature robustness was studied against segmentation variations, discretization schemes, etc. It was performed independent-to-outcome. |
| | Feature complementarity | ✓ | Extensively implemented, reducing 487 to 56 features. |
| **Model Assessment** | False-discovery correction | ✓ | Implementing Benjamini-Hochberg for univariate, AIC, and Fisher's methods for multivariate. |
| | Estimation of model performance | ✓ | The training model was further optimized on the validation set. |
| | Independent testing | ✓ | The independent test set was blind to training/validation. Feature selection was blind to outcome. |
| | Performance results consistency | ✓ | The consistency of results was demonstrated via 50 times randomly shuffling the {training-validation}/test sets. |
| | Comparison to conventional metrics | ✓ | Radiomics results were compared against conventional (clinical) metrics |
| | Multivariable analysis with non-radiomic variables | ✓ | Multivariate analysis of combined radiomics + clinical features were provided |
| **Clinical implications** | Biological correlate | ✓ | We provide intuition explaining why specific radiomics features (e.g. GLSZM-small zone large GL |

| | | | |
|---|---|---|---|
| | | | emphasis) appear in the multivariate model fit |
| | Potential clinical application | ✓ | Potential clinical application is to predict/stratify patients' CAC score based on radiomics of MPSS |
| **Material availability** | Open data | ✗ | Not available |
| | Open code | ✓ | Provided in the Supplementary Materials section |
| | Open models | ✓ | The result of our independent-to-outcome feature selection process has been provided for future studies in radiomics of MPSS. |
